# Ex vivo hypothermic oxygenated perfusion allows extended heart preservation in a donation-after-circulatory-death porcine model

**DOI:** 10.1101/2025.10.09.25337185

**Authors:** Morgan K. Moroi, Yaagnik Kosuri, Cary Karcher, Anthony Campbell, Arianna Adamo, Diana Albino, Emre Bektik, Christine Chan, Kenmond Fung, Miroslav Sekulic, Shaheer K. Faruqi, Craig J. Goergen, Melissa Tamimi, Koji Takeda, Giovanni Ferrari

## Abstract

**Objectives:** Donation after circulatory death (DCD) expands the donor heart pool but is limited by warm ischemia and short preservation times. Hypothermic oxygenated perfusion (HOPE) extends storage beyond the 4–6-hour limit of static cold storage (SCS), yet cellular and molecular responses remain undefined. We evaluate cardiomyocyte integrity and functional recovery of DCD porcine hearts after in situ reanimation with normothermic regional perfusion (NRP) and preservation by SCS (2h) or HOPE (24h) and assess the impact of NRP under DCD conditions.

**Methods:** Nine Yorkshire pigs underwent DCD cardiectomy. Six animals experienced 15 min warm ischemia followed by 60 min NRP. Three hearts were preserved for 2h with SCS and three for 24h with HOPE. A second DCD group (n=3) underwent direct procurement without NRP and 2h of HOPE preservation. All hearts were reanimated by normothermic machine perfusion to assess rhythm and contractility. Cardiomyocyte viability, transcriptomics, and metabolomics were analyzed.

**Results:** After DCD+NRP, 2h SCS preserved intact cardiomyocyte viability. HOPE maintained measurable, though reduced, viability at 24h, while 24h SCS failed even under donation after brain death (DBD) conditions. Transcriptomic and metabolomic analyses showed marginal differences between 2h SCS and 24h HOPE. All 24h HOPE hearts regained sinus rhythm but showed reduced contractility vs 2h SCS. Without NRP, 2h HOPE hearts showed the lowest viability and contractility.

**Conclusions:** HOPE supports extended preservation of DCD hearts, but viability and function decline by 24h. NRP is essential for functional recovery of hearts preserved at hypothermic temperatures in a porcine preclinical DCD model.

**Graphical Abstract:** 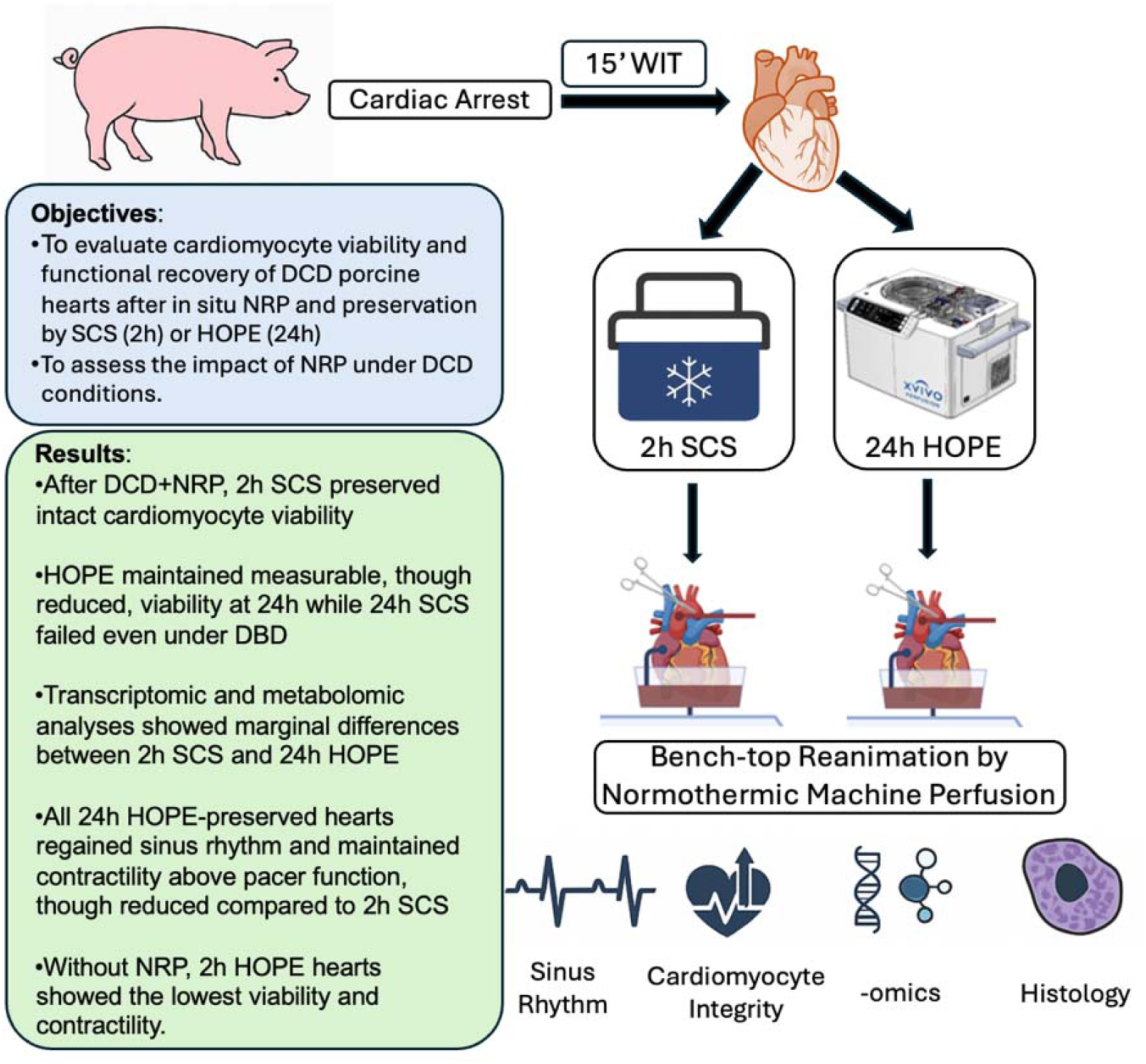

## Introduction

Cardiac transplantation is the gold standard for end-stage heart failure, yet the supply of suitable donor hearts has not kept pace with rising demand, creating an urgent need to expand the donor pool^1,2,3^. Expanding donor criteria is one potential solution to address this shortage. While donation after brain death (DBD) remains the standard source of donor hearts, donation after circulatory death (DCD) has emerged as a promising but underutilized option^4,5,6^. Two procurement strategies are currently applied clinically for DCD hearts. Both begin with withdrawal of life support, cardiac arrest confirmed by asystole, and a mandated stand off period of several minutes^7^ ^8^ ^9^. In the first method, Direct Procurement Perfusion (DPP), uses a normothermic physiological perfusion systems such as the OCS TransMedics Device^4^. The heart is immediately protected by delivery of antegrade cardioplegia before it is excised and reanimated within a normothermic machine perfusion (NMP) device. The second technique, thoracoabdominal normothermic regional perfusion (TA-NRP),vreferred as NRP in this work, requires that the donor chest be opened, central cannulation established, and the heart reanimated in situ via a cardiopulmonary bypass or extracorporeal membrane oxygenation circuit for evaluation before arrest and cardiectomy. This strategy culminates in the successful weaning of mechanical support and allows for functional assessment. Recently, a group from Duke University transplanted a pediatric heart following DCD procurement with the inclusion of a bench-top NMP circuit^11^. The NMP circuit allows for reperfusion, resumption of normal sinus rhythm and contractility ex-situ, though the heart is not physiologically loaded^12^ ^13^ ^14^ ^15^.

In addition to procurement strategies, preservation methods remain a critical barrier. Static cold storage (SCS) is used for cardiac allografts, but its upper limit is 4 to 6 hours, and prolonged ischemia correlates with primary graft dysfunction (PGD)^1,16,17,18^. Hypothermic oxygenated perfusion (HOPE) offers an alternative reducing ischemic injury during storage^19^. XVIVO’s HOPE device has recently shown encouraging results in clinical DBD trials^19^. In a recent study, we reported that HOPE, under cardio protected DBD conditions, enables 24h *ex vivo* heart preservation with cardiomyocyte integrity, normal gross and microscopic architecture, and rapid functional recovery on bench-top reperfusion^20^.

This new study aims to evaluate cardiomyocyte viability and functional recovery of DCD porcine hearts after in situ reanimation with NRP and preservation by SCS (2h) or HOPE (24h), and to assess the impact of NRP by comparing DCD hearts procured with or without NRP (referred as Direct Procurement, DP, in this work).

## Methods

Detailed protocols can be found in the Supplemental Materials and Methods section.

### Animal characteristics

Female American Yorkshire pigs (n=9, 40-50 kg, 4-6 months old) were used: SCS vs HOPE extended preservation and TA-NRP vs direct procurement. This study was approved by the Columbia University (IACUC #AC-AABX9650) and performed under the NIH Guide for the Care and Use of Laboratory Animals.

### Simulating DCD Procurement model

Swine donors underwent anesthesia with Telazol, propofol, and buprenorphine, were intubated, and maintained with isoflurane, midazolam, and propofol. After sternotomy, the great vessels were exposed and systemic heparinization achieved. Venous, arterial, and cardioplegia cannulae were placed. For DCD with NRP (n=6), animals received vecuronium, ventilation was withdrawn, and functional warm ischemia began at MAP <50 mmHg. After asystole and a 15min no-touch period, NRP was established, hearts defibrillated if needed, and reperfused for 1h before arrest with cold cardioplegia and cardiectomy. For direct procurement (n=3), hearts were arrested after 15min warm ischemia without NRP.

### Preservation

SCS hearts (n=3) were stored in 1L of XVIVO Heart Solution at 4°C for 2h. HOPE hearts were preserved in the XVIVO Heart Assist Transport device for 2h (n=3) or 24h (n=3) at 8°C with carbogen (95%O₂/5% CO₂). The circuit was primed with XVIVO cardioplegia plus red blood cells, supplement, insulin, antibiotics, heparin, and potassium. Hearts were connected via an aortic cannula, LV vented with a mitral shunt, and perfused at 20mmHg. Heart weight was recorded at the end of preservation.

### Bench-Top Normothermic Machine Perfusion

After preservation, nine hearts (n=9) were reanimated on a custom NMP circuit for 2h to model ischemia–reperfusion and assess rhythm and contractility under unloaded conditions. The circuit included a venous reservoir, oxygenator, heater–cooler, and roller pump, and was primed with 500mL autologous blood plus heparin. Hearts were cannulated via the ascending aorta, LV vented through the mitral valve, and atria opened for venous return. After de-airing, reperfusion was initiated and hearts paced at 100bpm with epicardial electrodes.

### Myocardial Motion Video Quantification

A custom Python pipeline (Python 3.10; OpenCV, NumPy, SciPy) was used to quantify myocardial motion during NMP reanimation. Videos were calibrated using a pixel-to-centimeter scale, and a region of interest was defined over the anterior ventricles. To correct for camera movement, frames were stabilized with dense optical flow before recomputing motion vectors. Velocity traces were derived from the mean vector magnitude within the ROI, converted to mm/s using calibration and frame rate, and smoothed with a Savitzky–Golay filter. Systolic and diastolic phases were tracked, and complete cardiac cycles were analyzed for motion quantification.

### Histological analysis

Endomyocardial biopsies (RV septum, LV free wall, apex) were collected after 2h of NMP. Samples were fixed in 10% formalin, paraffin-embedded, sectioned (5µm), and stained with H&E, Masson’s trichrome, and TUNEL. Slides were examined for coagulative myocyte necrosis, inflammation, edema, hemorrhage, fibrosis, thrombosis, and apoptosis.

### Flow Cytometry

RV septum biopsies (∼0.2–0.3 g) were enzymatically digested filtered, and RBC-lysed. Cells were fixed, permeabilized, and stained with Alexa Fluor 647 anti-cardiac troponin T and DAPI. Intact cardiomyocytes were identified as troponin⁺/DAPI^+^ and gated by FSC/SSC to exclude debris and apoptotic bodies. Cardiomyocyte size and troponin expression were used as indicators of preserved cellular integrity and viability.

### RNA Extraction and Bulk RNA sequencing Analysis

RNA was extracted from apex biopsies (RNeasy Mini Kit, Qiagen), quality-checked (RIN >6, Bioanalyzer), and sequenced (Illumina 2× 75bp, TruSeq Stranded mRNA kit, Azenta Life Sciences). Reads were trimmed (Trimmomatic), aligned to the pig genome (STAR), and counted (featureCounts). Differential expression was performed with DESeq2 (padj<0.05, |log₂FC| >1). Results were visualized with volcano plots and heatmaps (DataMap), and KEGG pathway enrichment was assessed (ShinyGO).

### Untargeted Metabolomic Profiling

Global untargeted metabolomics was performed on flash-frozen left ventricular biopsies using UHPLC–high resolution accurate mass spectrometry.

Metabolites were extracted by protein precipitation with methanol:acetonitrile (1:1) and analyzed using both hydrophilic interaction chromatography (HILIC) and reverse-phase separation in positive and negative ion modes. Compounds were identified using mzVault, mzCloud, ChemSpider, and public metabolite databases (HMDB, KEGG, LipidMAPS). Differential metabolites were determined by Student’s t-test with reporting of p-value, q-value, and fold-change.

### Statistics

For comparison of two groups, unpaired two-tailed Student’s t-test followed by two-tailed Mann–Whitney U test was used.

## Results

### Study Design

This study evaluated extended hypothermic oxygenated perfusion (HOPE) for donation after circulatory death (DCD) heart preservation and the role of normothermic regional perfusion (NRP). Nine Yorkshire pigs were studied. Six underwent simulated DCD with 15min warm ischemia (**Supplemental video 1**) and 60 min NRP (**Supplemental video 2**); three hearts were preserved 2h by static cold storage (SCS) and three for 24h by HOPE. A second group (n=3) underwent direct procurement without NRP and 2h HOPE preservation. All grafts were reanimated on a normothermic machine perfusion circuit, with outcomes including cardiomyocyte viability, RNA sequencing, metabolomics, and contractility assessment (**Supplemental Fig 1**).

### Functional recovery of HOPE-preserved DCD hearts following bench-top normothermic reperfusion

Functional recovery was assessed using a custom bench-top normothermic machine perfusion (NMP) system following hypothermic preservation (**Fig 1A**). Hearts preserved for 2h with SCS after DCD procurement and 1h of NRP consistently regained sinus rhythm and demonstrated robust contractile motion (**Supplemental Video 3**). By contrast, hearts preserved for 24h with HOPE, which is three times the current clinical usage, showed a reduction in contractile amplitude (**Supplemental Video 4**). Despite this reduction, all HOPE- preserved grafts were able to restart, regain sinus rhythm, and maintain contractility above intrinsic pacer function throughout reperfusion. This finding contrasts with SCS preservation at 24h, which under favorable DBD conditions failed to reanimate (**Fig 1B** and **Supplemental Video 5**). Quantitative motion analysis confirmed significant differences in contractile velocity between the two groups. Average myocardial motion magnitude in the HOPE 24h group was reduced compared with the SCS 2h group indicating partial preservation of function (**Fig 1C** and **1D**).

**Figure 1.**
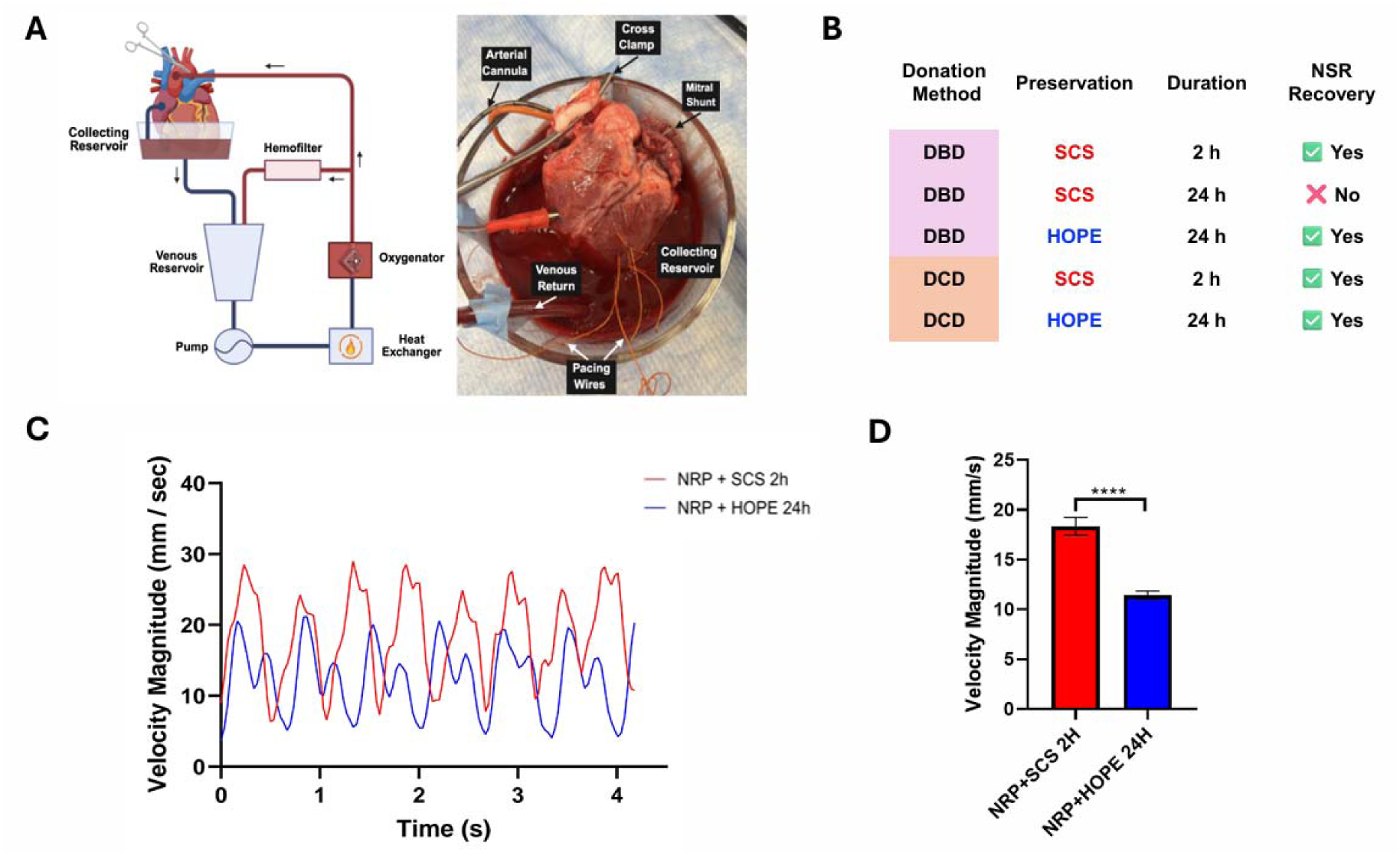
Functional recovery of DCD hearts following preservation and bench-top normothermic reperfusion. (A) Schematic and photograph of the normothermic machine perfusion (NMP) circuit. (B) Experimental groups: DBD and DCD hearts preserved by SCS or HOPE for 2h or 24h. (C) Velocity traces show greater contractile amplitude in NRP + SCS 2h vs NRP + HOPE 24h. (D) Average velocity magnitude (mm/s) ± SEM confirmed higher activity in NRP + SCS 2h (****p < 0.0001, Mann–Whitney U).

### Gross examination and Histopathological analysis of porcine hearts preserved with either static cold storage or HOPE after DCD Procurement

Across groups, hearts gained weight during preservation and reperfusion, with HOPE-preserved grafts showing modest increases post-preservation, while all groups reached ∼15–20% of baseline after NMP (**Supplemental Fig. 2**). Following completion of the NMP circuit, ventricles were cut open; there was no evidence of necrosis in the NRP+SCS 2h group. In contrast, in the NRP+HOPE 24h hearts there was visibly trace evidence of necrosis within the myocardium (**Fig 2A**). In our previous works evaluating DBD hearts stored for 24 h on HOPE, no necrosis was observed (^20^). Histological evaluation was performed on endomyocardial biopsies collected from the left and right ventricles after DCD procurement and preservation by either SCS or HOPE. Across all groups, H&E and trichrome staining showed preserved overall myocardial architecture without evidence of interstitial fibrosis or thrombosis. Edema was a consistent finding, present in all samples and was more pronounced in HOPE-preserved hearts (24h) (**Fig 2B** and **C; Supplemental Fig 3A**), consistent with our previous work (^20^). Coagulative myocyte necrosis (CMN) was very rare, identified in only one NRP heart preserved in HOPE for 24h. Inflammation was minimal, limited to focal neutrophil infiltrations in one specimen. No cases demonstrated interstitial hemorrhage or vascular thrombosis. TUNEL staining revealed scattered apoptotic cardiomyocytes and interstitial cells in both SCS- and HOPE-preserved hearts, consistent with sub-lethal injury following ischemia-reperfusion (**Fig 2D**). However, no diffuse TUNEL positivity or confluent necrosis was observed, underscoring the absence of widespread irreversible cell death under either condition. Taken together, these findings align with functional and flow cytometric analyses, where HOPE preserved measurable viability despite evidence of structural stress.

**Figure 2.**
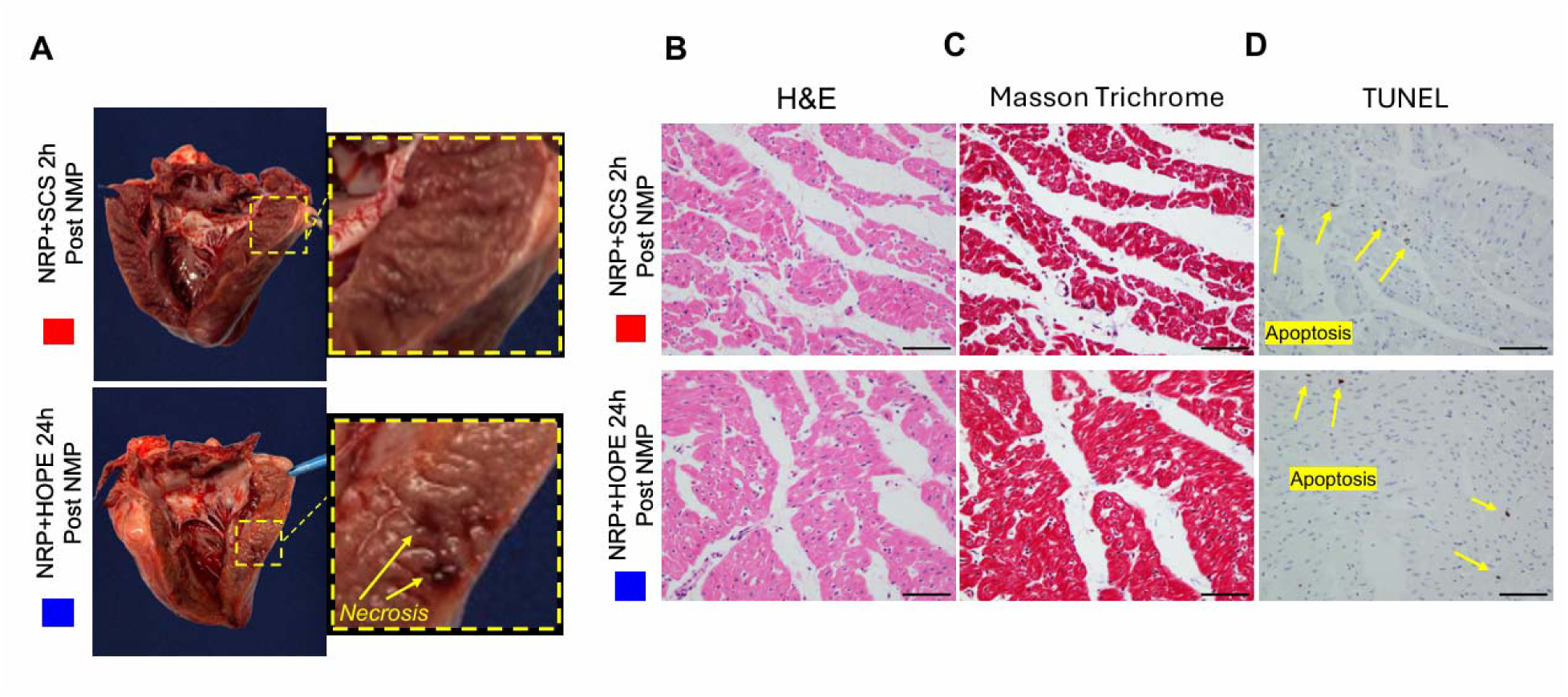
Gross morphology and histology of swine hearts after NMP. (A) Hearts after reanimation by NMP following 2h SCS (red) or 24h HOPE (blue). Whole heart views with opened left ventricle show focal necrosis in 24h HOPE, absent in 2h SCS. (B–C) H&E and Masson Trichrome of the right ventricular septum show preserved architecture across groups. (D) TUNEL staining of the right ventricular septum shows comparable apoptotic cell positivity between groups. Scale bar: 100 µm.

### Reduced but preserved cardiomyocyte integrity after 24h of Hypothermic oxygenated preservation conditions

Flow cytometric analysis was performed to quantify cardiomyocyte integrity following preservation under different procurement strategies. DAPI and anti-cardiac troponin T double-positive events (P1 gate) were further filtered by gating for SSC-A and FSC-A to exclude debris. After DCD procurement with NRP and 2h of SCS, the population of isolated cardiomyocytes maintained cellular integrity and viability (P2 gate). Extending preservation under HOPE to 24h reduced overall viability compared with 2h SCS but a measurable population of intact cardiomyocytes was retained, demonstrating preserved cellular integrity despite prolonged storage and supporting the functional recovery (**Fig. 3A** and **B**). This finding contrasts with SCS at 24h, which under favorable DBD conditions failed to yield any viable cardiomyocytes, consistent with irreversible cellular injury (**Supplemental Fig 4**).

**Figure 3.**
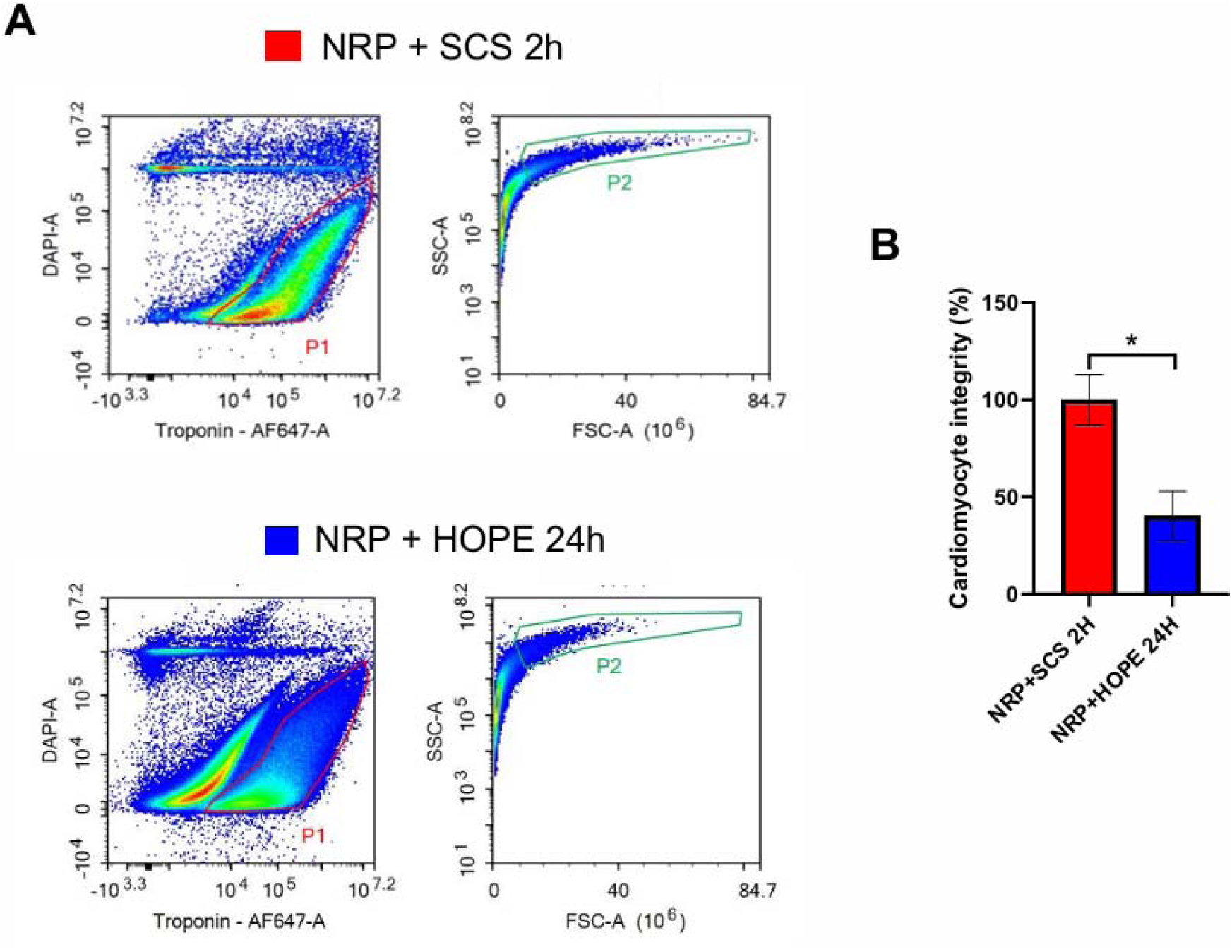
Cardiomyocyte integrity after extended HOPE preservation. (A) Flow cytometry of isolated cardiomyocytes after NRP + SCS 2h (top) or NRP + HOPE 24h (bottom). (B) Quantification of troponin⁺/DAPI⁺ cardiomyocytes (P2 gate) showed significantly lower viability in NRP + HOPE 24h vs NRP + SCS 2h (*p < 0.05, Mann–Whitney U). Data are mean ± SEM.

### Transcriptomic and Metabolomics analysis of Extended Hypothermic oxygenated preservation under DCD conditions

To investigate the cellular and molecular consequences of extended hypothermic oxygenated preservation, transcriptomic and metabolomic profiling was performed on DCD hearts preserved for 2h with SCS or 24h with HOPE. RNA sequencing revealed minimal transcriptional differences between groups. Differential gene expression analysis identified only a limited number of genes meeting statistical significance (p < 0.05, |log2FC| > 1), with limited enrichment of gene ontology terms across biological processes, molecular functions, or cellular components. Pathway-level interrogation confirmed the absence of coordinated transcriptional shifts, indicating that extended HOPE preservation maintained a transcriptomic state broadly comparable to short-term SCS (**Fig 4A** and **B** and **Supplemental Fig 5A-C**). KEGG pathway enrichment analyses of upregulated DEGs shows that mostly inflammation-related signaling reached statistical significance (FDR > 0.01) while KEGG pathways for downregulated DEGs did not reach significance (**Fig 4C**). Data are accessible at GEO #[*<u>RNA Sequencing Analysis</u>*]. Metabolomic profiling demonstrated molecular stability across preservation strategies. While 33 metabolites were found to be differentially abundant (p < 0.05, |log2FC| > 0.693), pathway analysis revealed no consistent alterations in core metabolic networks. Global clustering of annotated metabolites showed substantial overlap between the two groups, underscoring the preservation of metabolic homeostasis during prolonged HOPE storage (**Fig 4D** and **E** and **Supplemental Table 1**). Data are accessible at GEO #<u>[Metabolomics Analysis</u>].

**Figure 4.**
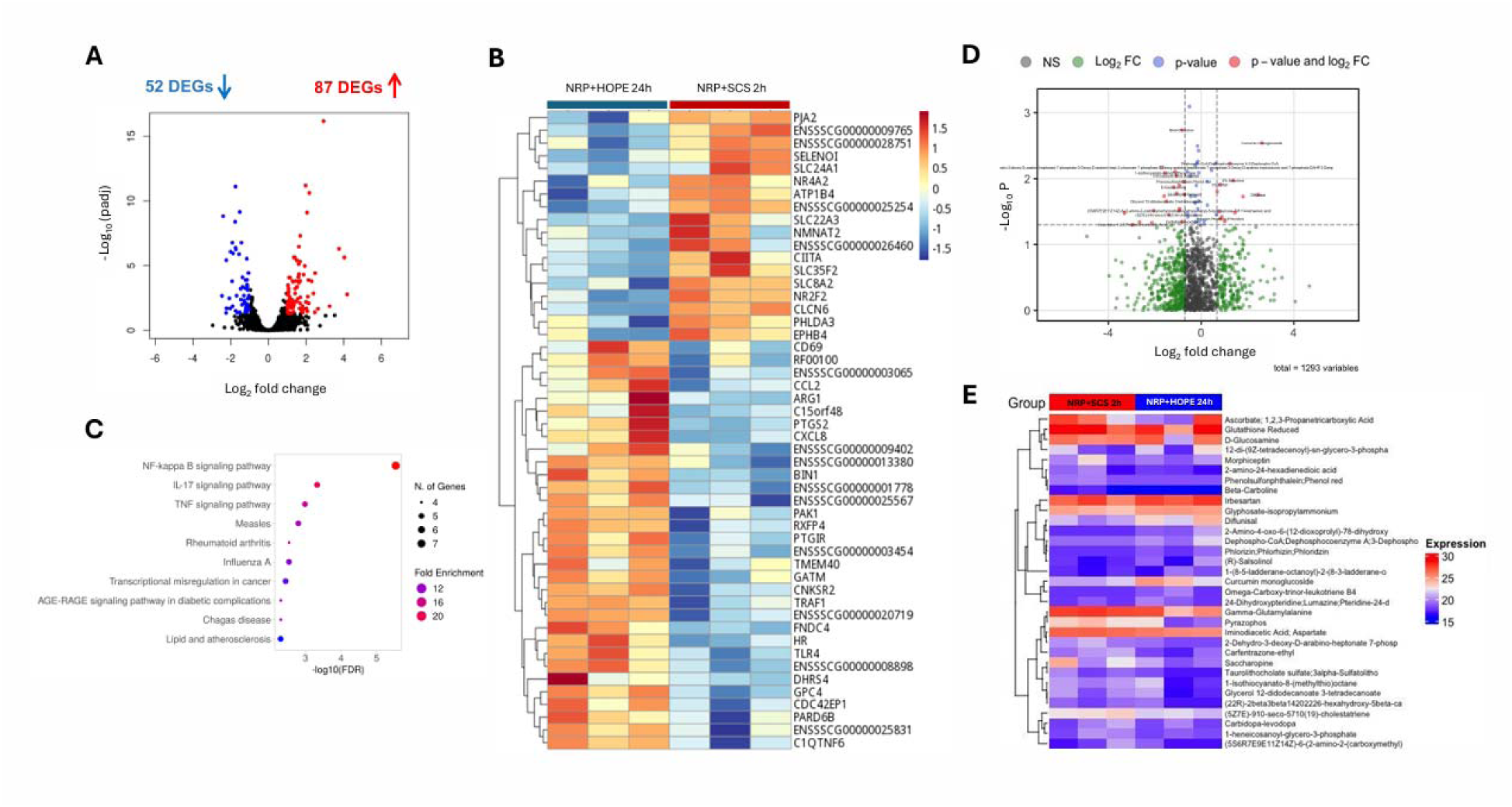
Transcriptomic and metabolomic profiling of DCD hearts. (A) Volcano plot of differentially expressed genes (DEGs) between NRP + HOPE 24h and NRP + SCS 2h. (B) Heatmap of top 50 DEGs shows distinct expression patterns. (C) KEGG enrichment of upregulated DEGs highlights inflammation-related pathways (FDR < 0.01). (D) Volcano plot of metabolomic analysis shows significantly altered metabolites (red). (E) Supervised clustering heatmap of 33 significant metabolites shows no consistent pathway-level shifts.

### Impact of NRP in DCD heart procurement: functional recovery, cardiomyocyte integrity and histopathological analysis

To determine whether in situ reanimation with NRP influences porcine heart physiology during DCD procurement, we compared hearts preserved after NRP, 2h SCS, and 2h of normothermic benchtop reanimation (NRP + SCS 2h), the clinical standard, versus no NRP, 2h HOPE (HOPE 2h) followed by 2h of normothermic benchtop reanimation in both groups. Hearts procured without NRP exhibited the most severely impaired functional recovery, with depressed myocardial motion and reduced ability to sustain contractility above intrinsic pacer function (**Fig. 5A** and **B; Supplemental Video 6**). Assessment of cardiomyocyte integrity confirmed these findings. Hearts preserved with HOPE 2 h in the absence of NRP displayed the lowest proportion of viable troponin-positive, DAPI-positive cardiomyocytes compared with both NRP + SCS 2h and NRP + HOPE 24h. (**Fig. 5C** and **DB**). These results suggest that the absence of NRP has a detrimental effect on the viability of myocardial tissue in DCD-procured hearts and that NRP may be critical for stabilizing DCD hearts prior to preservation in a porcine model. The histological evaluation of endomyocardial biopsies stained with H&E and Masson Trichrome revealed preserved general myocardial architecture of the right ventricular septum and left ventricular free wall. Edema was consistently observed in all samples, while no signs of fibrosis, thrombosis, interstitial hemorrhage and coagulative myocyte necrosis were reported. Fragmented DNA was identified by TUNEL staining in scattered cardiomyocytes and interstitial cells (dark brown), though not consistent with a diffuse apoptotic pattern. (**Supplemental Fig 3A** and **B**).

**Figure 5.**
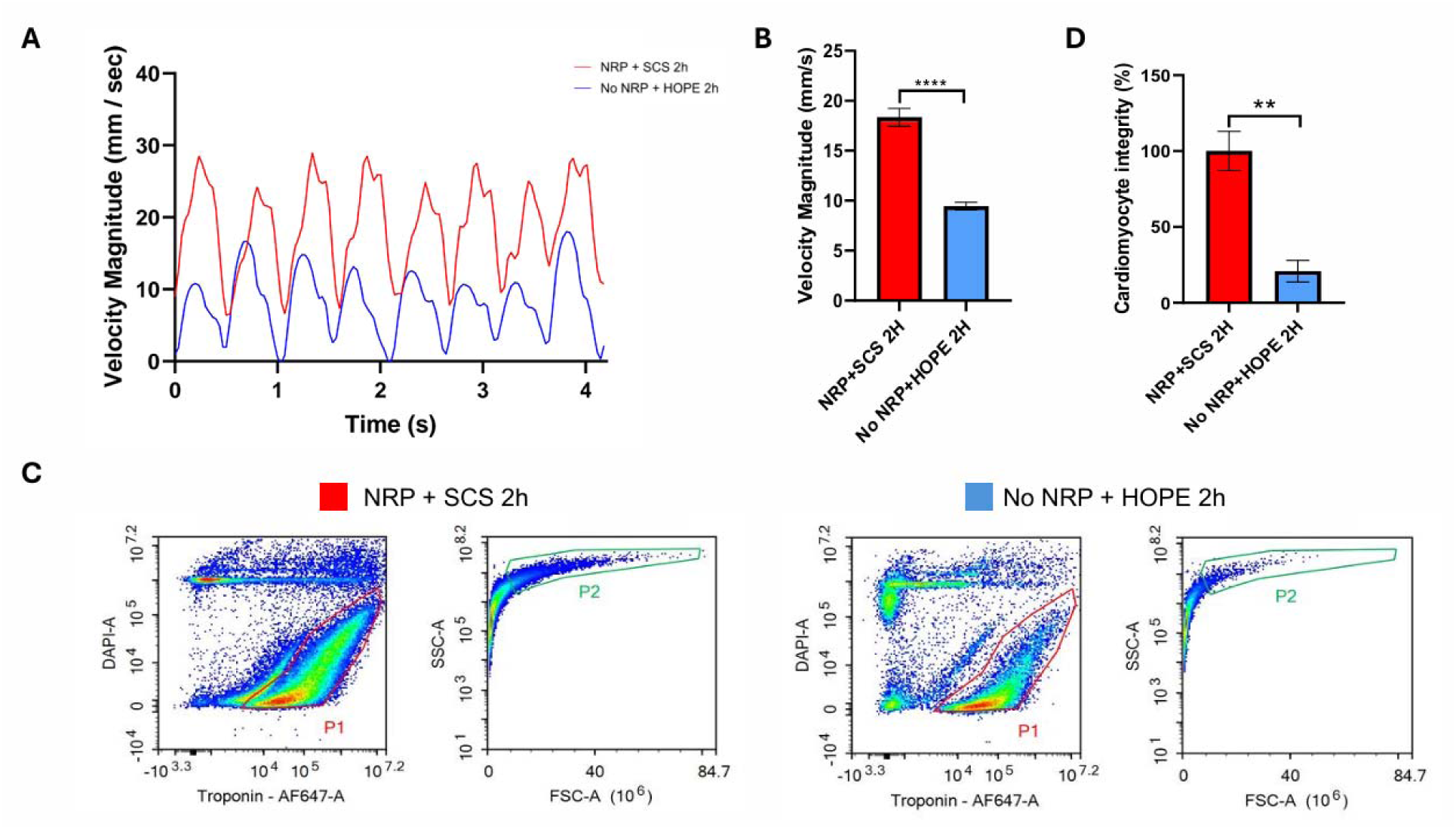
Impact of NRP in DCD hearts procurement: functional recovery and cardiomyocyte integrity. (A) Velocity traces during bench-top reperfusion show higher contractile activity in NRP + SCS 2h vs No NRP + HOPE 2h.c(B) Quantification confirmed significantly higher contractility in NRP + SCS 2h (****p < 0.0001, Mann–Whitney U). Data are mean ± SEM.c(C) Flow cytometry plots of troponin/DAPI staining show greater viability in NRP + SCS 2h. (D) Quantification of troponin⁺/DAPI⁺ cardiomyocytes (P2) confirmed significantly higher viability in NRP + SCS 2h (**p < 0.01, Mann–Whitney U). Data are mean ± SEM.

### Impact of NRP in DCD hearts procurement: Transcriptomic and Metabolomics analysis

Transcriptomics: Bulk RNA sequencing revealed minimal differences between the two groups. Although a limited number of individual genes met nominal significance, KEGG pathway analysis failed to identify consistent changes (FDR>0.01). Unsupervised clustering confirmed substantial overlap in the global transcriptomic profiles, indicating that surviving cardiomyocytes retained comparable transcriptional signatures regardless of NRP exposure (**Fig. 6A** and **B; Supplemental Fig 5D-F**). Metabolomics: Similarly, metabolomic profiling identified 40 differentially abundant compounds (p<0.05, |log2FC| >0.693), but these differences did not converge into coherent shifts in central energy metabolism, amino acid pathways, or oxidative stress networks (**Supplemental Table 2**). Both supervised and unsupervised clustering analyses showed significant overlap between groups, suggesting metabolic stability independent of procurement strategy (**Fig 6C** and **6D**). Taken together, these findings indicate that the benefits of NRP observed at the cellular and functional levels are not explained by broad transcriptional or metabolic reprogramming. Data are accessible at GEO #[*<u>RNA</u> <u>Sequencing Analysis</u>*] and GEO #[<u>Metabolomics Analysis</u>].

**Figure 6.**
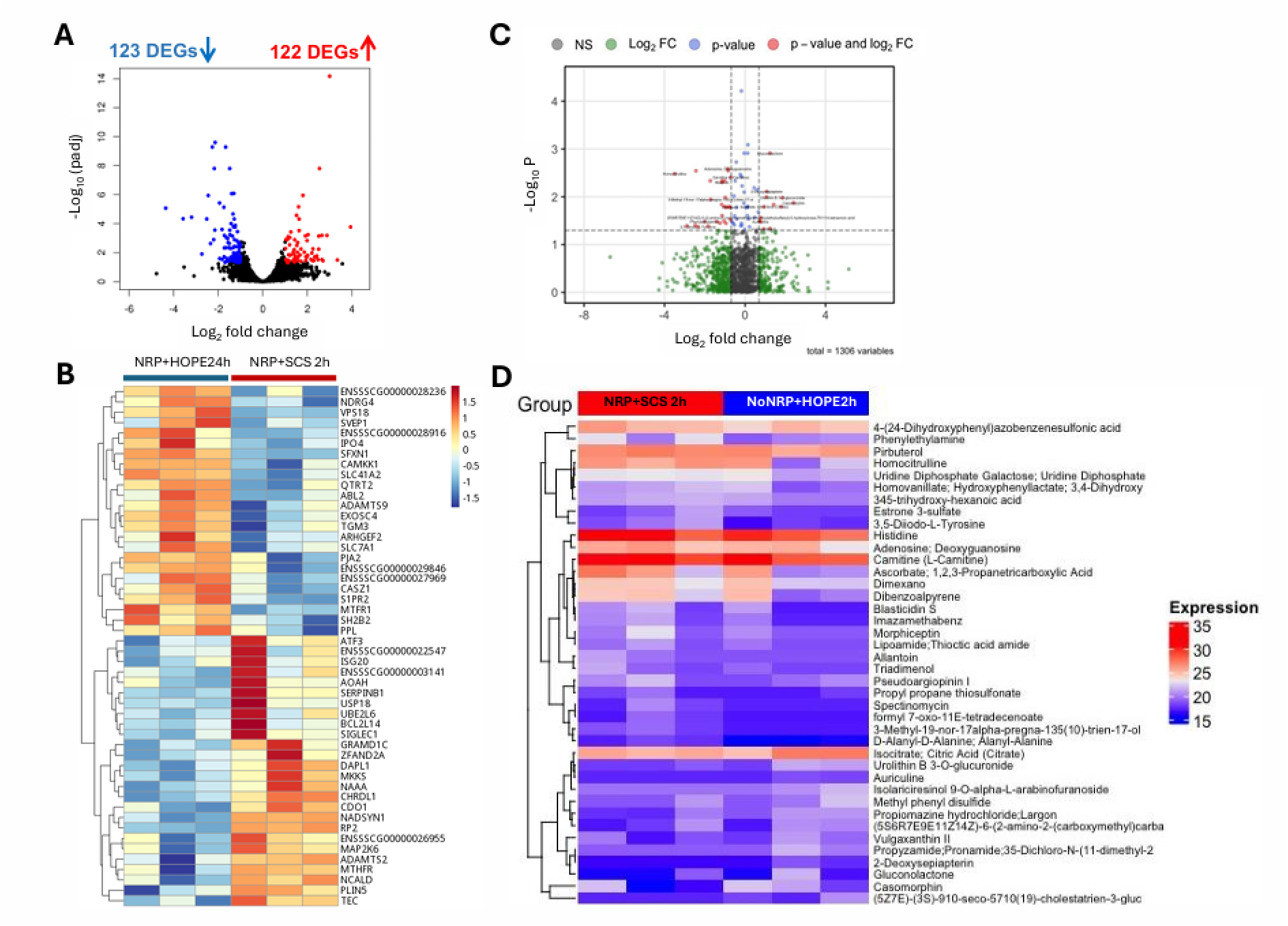
Transcriptomic and metabolomic profiling of DCD hearts preserved with NRP + SCS (2h) or No NRP + HOPE (2h). (A) Volcano plot shows 245 differentially expressed genes (122 upregulated, 123 downregulated). (B) Heatmap of the top 50 DEGs demonstrates distinct transcriptional patterns. (C) Volcano plot of metabolomics highlights significantly altered metabolites (red). (D) Heatmap of 40 significant metabolites (p<0.05, |log₂FC| >0.693) shows group separation without consistent pathway-level enrichment.

## Discussion

In this study, we employed a porcine model to evaluate whether hypothermic oxygenated perfusion (HOPE) could extend preservation of DCD hearts, and to better characterize the role of normothermic regional perfusion (NRP) in enabling functional recovery.

Following DCD procurement with 15 minutes of warm ischemia and 60 minutes of NRP, hearts preserved for 2h in SCS retained intact cardiomyocyte viability and demonstrated robust contractile recovery on bench-top reperfusion. In contrast, when preservation was extended to 24h, even in the most favorable DBD protocol, SCS-stored hearts resulted in complete loss of viable cardiomyocytes. By comparison, HOPE maintained measurable cardiomyocyte viability and enabled hearts to restart, regain sinus rhythm, and sustain contractile activity of DCD- procured hearts after 24h, more than six times the current clinical usage. Although contractility was significantly reduced compared with short-term SCS, the ability to achieve partial functional recovery highlights the capacity of HOPE to support extended preservation within the more challenging DCD setting, in which warm ischemia and circulatory arrest present additional barriers. While promising, the declining contractility and cardiomyocyte viability indicate that utilization of organs preserved for 24h in this condition may not be yet optimal for transplantation experiments.

To explore the mechanisms underlying partial functional recovery after 24 h of HOPE, we assessed cardiomyocyte viability, RNA sequencing, and metabolomic profiles. Flow cytometry revealed that while cardiomyocyte integrity was reduced after extended HOPE compared with 2h SCS, a significant fraction of troponin-positive, DAPI-positive cells was retained, in contrast to the complete loss observed with 24h SCS under DBD. This preservation of viable cells aligns with the ability of HOPE hearts to restart and maintain sinus rhythm despite reduced contractility. Interestingly, transcriptomic and metabolomic analyses revealed only marginal differences between 2h SCS and 24h HOPE. RNA sequencing identified a relatively small number of differentially expressed genes, with moderate enrichment of inflammation-related pathways, which is often observed in ex vivo perfused hearts and may be a potential improvement point in prolonged heart preservation. Similarly, metabolomic profiling detected 33 differentially abundant compounds, but pathway analysis failed to reveal consistent alterations in central metabolic networks. These findings suggest that although cardiomyocyte integrity is reduced at the structural level, the surviving cells maintain essential transcriptional and metabolic programs. Interventions aimed at stabilizing mitochondria, or preventing edema could synergize with HOPE to further extend functional preservation. It also highlights the limitation of bulk RNA-seq and metabolomics: these methods primarily capture surviving cell populations and may underestimate injury severity in the total myocardium.

Our second finding is that NRP may be critical for safe organ harvest and enabling functional recovery following DCD procurement. Hearts procured without NRP exhibited the lowest cardiomyocyte integrity and most depressed functional recovery. Flow cytometry confirmed greater cellular injury in the no-NRP group, with significant decline in cardiomyocytes recovery. Transcriptomic and metabolomic analyses revealed minimal differences between NRP and no- NRP groups. We can propose several mechanisms by which NRP exerts its benefit: restoration of ATP levels before cardioplegia, washout of accumulated metabolites and microthrombi, stabilization of mitochondrial function, and preconditioning against reperfusion injury.

Our results suggest that NRP provides essential resuscitation and stabilization, enabling subsequent preservation by HOPE to achieve meaningful functional recovery. Together, these findings support a translational strategy combining NRP and HOPE for DCD heart preservation. NRP appears essential for initial stabilization, while HOPE provides a platform for extending preservation beyond the limits of SCS.

Several limitations warrant discussion. First, the sample size was small, reflecting the complexity of large animal models. Second, the porcine model, while physiologically relevant, may not fully recapitulate human clinical DCD conditions, including variable warm ischemia times and comorbidities. Third, functional recovery was assessed ex vivo using NMP rather than orthotopic transplantation. Our optical-flow contractility analysis, based on Farnebäck dense optical flow, was intended as an objective and reproducible supplement to visual rhythm recovery and electrophysiology activity, but it only estimates velocity magnitude of the anterior ventricular wall and cannot fully capture global myocardial function. Moreover, the heart contractility improves along with the NMP circuit duration, however, in this study, the video acquisition sampling time was not standardized along all the experiments. Fourth, as discussed, bulk RNA-seq and metabolomics are limited by survivor bias and may underestimate injury in the total myocardium.

## Conclusion

HOPE enables extended preservation of DCD hearts with preserved molecular programs and sustained contractile activity, though reduced viability and function suggest the upper limit of utilization in this model may be less than 24h. NRP appears essential for procurement under DCD conditions, supporting its role in achieving functional recovery in preclinical transplantation studies in a porcine model.

## Data Availability

All data produced in the present study are available upon reasonable request to the authors

## Acknowledgement

The Molecular Pathology Shared Resource of the Herbert Irving Comprehensive Cancer Center (#NIH P30 CA013696); Sulzberger Columbia Genome Center (NIH P30CA013696)

## Author contributions

Conceptualization: GF,MKM,YK

Methodology: MKM,YK,CK,AA,DA,EB,CG,KT

Investigation: MKM,YK,CK,AC,AA,DA,EB,CH,KF,MS,SF,CG,MT,KT,GF

Visualization: MKM,YK,CK,AA,DA,EB,SF,GF

Funding acquisition: GF

Writing, original draft: GF,YK,MKM,CK,DA

Writing, review & editing: MKM,YK,CK,AC,AA,DA,EB,CH,KF,MS,SF,CG,MT,KT,GF

## Data and Material Availability

RNA sequencing and metabolomic datasets are available at GEO repository [<u>*RNA Sequencing Analysis*</u>; <u>Metabolomics Analysis</u>].

## SUPPLEMENTAL MATERIALS

### Supplemental Materials and Methods

#### Preservation

The hearts selected for SCS clinical control (n=3) were stored in 1L of XVIVO Heart Solution at 4 °C for 2 hours. Hearts randomly selected for HOPE preservation were stored in the XVIVO Heart Assist Transport (XVIVO Group, Gothenburg, Sweden) for 2 hours (n=3) or 24 hours (n=3). HOPE provides oxygenated perfusion using carbogen gas (95% O2, 5% CO2) at 8 °C. The heart preservation device was primed with XVIVO cardioplegic solution supplemented with 350ml packed red blood cells, 3mL of XVIVO Heart Solution Supplement, 20 IU insulin (Humulin ELN-1325240), 50 mg Imipenem and Cilastatin (WG Critical Care 44567– 705–10, 5000 IU unfractionated heparin (Sagent Pharmaceuticals 25021-0400-30) and 10 mmol Potassium (Hospira 00409-6635-01). The hearts are connected to the device through an aortic cannula and the tubing is deaired. A silastic shunt is fastened across the mitral valve to vent the left ventricle, preventing overdistention. Perfusion within the device was carried out at an aortic root pressure of 20 mmHg for 2 hours or 24 hours. Heart weight was measured at the end of preservation for all groups.

#### Donor Surgical Procedure: Simulating DCD Procurement model

The swine donor underwent anesthesia induction with intramuscular Telazol (6 mg/kg), propofol (1 mg/kg), and buprenorphine (0.01-0.03 mg/kg). Animals were endotracheally intubated in the prone position and rotated to a supine position for mechanical ventilation and surgery. Anesthesia and analgesia were maintained intraoperatively with inhaled isoflurane and/or continuous infusions of midazolam (0.2 mg/kg/hr) and propofol (3-5 mg/kg/hr). A midline sternotomy was performed with a bone saw, and the heart and great vessels were exposed. Intravenous heparin (300 IU/kg) was given for systemic anticoagulation. The right atrium was cannulated with a single-stage 24 Fr venous cannula (DLP Medtronic, Minneapolis, MN). The distal ascending aorta was cannulated with a one-piece 18 Fr arterial cannula (EOPA Medtronic, Minneapolis, MN). The aortic root was cannulated with an antegrade cardioplegia cannula (Surge Cardiovascular, Grand Rapids, MI).

Withdrawal phase for circulatory arrest: To replicate procurement following circulatory death criteria, the pigs (n=6) were first confirmed as deeply sedated and administered intravenous vecuronium 0.6mg/kg bolus followed by infusion at rate of 0.6mg/kg/hr. After achievement of chemical paralysis, the donor swine vitals were closely monitored until a MAP less than 50 mmHg was established; this was noted as functional warm ischemia time. When pulsatile measurements completely diminished from the arterial line, circulatory arrest was confirmed by asystole on the electrocardiogram. This began a 15-minute no touch period, after which, NRP was initiated using a CPB circuit. Cardiac function gradually resumed, and hearts which fibrillated were electrically defibrillated. All hearts regained sinus rhythm and weaned off cardiopulmonary bypass. After one hour of reperfusion, hearts were arrested in a standard brain-dead fashion. The left atrial appendage and IVC were opened to vent the heart, the aorta was cross clamped proximal to the aortic cannulation site, and the hearts were arrested via antegrade delivery of two 1L bags of XVIVO Heart Solution cardioplegia, 1ml/L Heart Solution Supplement, and 25mmol/L sodium bicarbonate (McKesson #239985) at 4 °C. Sterile slush was applied during arrest, and a standard cardiectomy was performed. To assess the role of NRP, circulatory death parameters were standardized to group 1, however the hearts were procured by direct procurement (n=3) without NRP. In group-2, the aorta was cross-clamped following 15 minutes of warm ischemic time and the hearts were arrested using the same supplemented solution as group-1. Standard cardiectomies were performed, and the hearts were weighed at baseline for all groups.

#### Bench-Top Normothermic Machine Perfusion

At the conclusion of preservation, nine hearts (n=9) were reanimated on a bench-top NMP circuit for 2 hours to induce ischemic reperfusion injury and assess gross cardiac rhythm and contractility by simulating reperfusion in unloaded conditions. Our NMP circuit consisted of a venous reservoir with Capiox FX05 oxygenator (Terumo Cardiovascular, Ann Arbor, MI), 3T Heater-Cooler system (LivaNova, London, UK), and Quantum 4-in. roller pump (Spectrum Medical, Gloucester, UK), and the circuit was primed with 500cc of whole blood from the donor pig and roughly 1000-2000 IU unfractionated heparin. The hearts were loaded onto the circuit through canulation of the ascending aorta (8Fr Bio-Medicus NextGen Pediatric Arterial Cannula, Medtronic, Minneapolis, MN) using the Seldinger technique. The left ventricle was vented through a silastic tube positioned across the mitral valve; this allows blood to drain into the venous return. The atria were widely opened, and the return from the heart was drained into a collecting reservoir. The circuit was slowly initiated by advancing the arterial line, the aorta was de-aired, and an aortic cross-clamp was applied to begin 2 hours of normothermic reperfusion. Hearts were paced at 100bpm by electrodes on the anterior epicardium of both ventricles.

#### Myocardial Motion Video Quantification

A custom Python pipeline (Python 3.10; OpenCV, NumPy, SciPy) was developed to quantify myocardial motion during NMP circuit reanimation. First, a pixel-to-centimeter scale factor was determined by manually selecting two points 1 cm apart on a representative video frame. This yielded a mm/pixel factor for each video. Next, a rectangular region of interest (ROI) was defined over the anterior epicardial surface of the heart ventricles to capture the maximal contracting area. To reduce artifacts of camera movement, videos were stabilized using dense optical flow, where median displacement vectors were applied to generate affine transformations that reduced global motion. Within the stabilized ROI, dense optical flow was recomputed between consecutive frames to estimate myocardial velocity. A single velocity trace was obtained using the spatial mean of vector magnitudes within the ROI for each frame, and then converted from pixels per frame to mm/s using the calibration factor and frame rate. The signal was smoothed using a Savitzky-Golay filter (window length = 11 frames, polynomial order = 2) to suppress noise while preserving physiological features. Systolic and diastolic phases were tracked, and all complete cardiac cycles were used for motion quantification.

#### Histological analysis

Endomyocardial biopsies were collected from the right ventricular (RV) septum and left ventricular (LV) free wall and apex using an 8-mm biopsy punch at the end of two hours of NMP. Additionally, aortic valve leaflets, mitral valve leaflets and aortic root were collected. Tissues were fixed in 10% formalin for 24 hours, then transferred to 70% ethanol solution and embedded in paraffin. 5µm-thick sections were cut using a microtome and mounted on high-adherence glass microscope slides. Sections were stained with hematoxylin and eosin (H&E) and Masson’s trichrome and imaged on a Leica 10X microscope. Standard TUNEL staining was also performed to identify DNA fragmentation and evaluate cell apoptosis. Histological slides were evaluated across all groups for presence of coagulative myocyte necrosis (CMN), inflammation, edema, interstitial hemorrhage, fibrosis, thrombosis, and apoptosis.

#### Flow Cytometry

RV septum biopsies (0.2-0.3 g) were obtained using an 8mm surgical punch and washed in PBS 1x (Corning). Tissues were then minced into ∼2mm^3^ chunks, transferred to a warm enzymatic solution of DMEM (Gibco) with 450 □U/ml Collagenase I (Worthington, LS004196), 60 □U/ml DNase I (Millipore, DN25) and 60 □U/ml hyaluronidase (Worthington, LS002592), and incubated in a rotating mixer (∼65/70 rpm) at 37 °C for 1 hour. The enzymatic digestion was stopped with HBB buffer (2% HI FBS (Gibco) and 0.2% BSA (Prometheus) in Hanks’ Balanced Salt solution (Sigma, H9269)) and the solution was strained through a 100μm strainer. The suspended cells were centrifuged at 350g for 5 min at 4 °C and the pellet was resuspended in ACK lysing buffer (Gibco, A10492) to lyse red blood cells. After 5 min of incubation at room temperature, cells were washed with 9 □ml of DMEM (Gibco) and centrifuged at 350g for 5 □min at 4 □°C.

Then, cells were incubated for 10 min on ice in blocking solution (PBS + 0.5% BSA), followed by a wash in PBS1x. Cells were fixed in 2% PFA for 20 min on ice and washed twice in PBS1x. After fixation, cells were permeabilized with 0.1% Triton in PBS 1x 0.5% BSA for 20 min at 4°C. Cells were stained with Alexa Fluor 647 mouse anti-cardiac troponin T (BD Bioscience – 565744) 1:50 for 1h on ice in blocking solution. DAPI (Invitrogen) was added (1:3000) and cells were analyzed with the NovoCyte Penteon Flow Cytometer Systems 5 Lasers (Agilent). Data were analyzed with NoveExpress 1.6.2 software. Detected events were gated to distinguish intact cardiomyocytes from debris. Specifically, cardiomyocytes were identified by first gating for DAPI and anti-cardiac troponin T staining; the selected population was then gated by SSC-A and FSC-A to separate the intact cells from troponin-positive debris and small apoptotic bodies, thus reducing the incidence of false positive events. Cardiomyocyte size and troponin expression were used as indicators of preserved cellular integrity and viability. The flow cytometry analysis was performed at the Columbia Stem Cell Initiative Flow Cytometry core facility at Columbia University Irving Medical Center.

#### RNA Extraction and Bulk RNA sequencing Analysis

RNA was extracted from apex of the hearts and purified using RNeasy Mini Kit (Qiagen, 74106) as per manufacturer guidelines. Purified RNA samples were processed and sequenced by Azenta Life Sciences (Burlington, MA, USA). Briefly, RNA integrity was assessed by Bioanalyzer (Agilent), and samples with RIN score > 6 were selected for library preparation. TruSeq Stranded mRNA kit was used to prepare cDNA libraries of poly-A enriched mRNAs, excluding rRNAs, and sequenced as 2x75 bp paired-end reads on Illumina platform. Sequence reads were trimmed to remove possible adapter sequences and nucleotides with poor quality using Trimmomatic (v.0.36). The trimmed reads were mapped to the Pig_ERCC reference genome available on ENSEMBL using the STAR aligner v.2.5.2b. The STAR aligner is a splice aligner that detects splice junctions and incorporates them to help align the entire read sequences. BAM files were generated as a result of this step. Unique gene hit counts were calculated by using featureCounts from the Subread package (v.1.5.2). The hit counts were summarized and reported using the geneid feature in the annotation file. Only unique reads that fell within exon regions were counted. If a strand-specific library preparation was performed, the reads were strand-specifically counted. After extraction of gene hit counts, the gene hit counts table was used for downstream differential expression analysis. Using DESeq2, a comparison of gene expression between the samples was performed. The Wald test was used to generate p-values and log2 fold changes. Genes with an adjusted p-value < 0.05 and absolute log2 fold change > 1 were called as differentially expressed genes for each comparison and visualized in volcano plots. Heatmaps of DEGs were generated using DataMap (v0.11). KEGG pathways were analyzed on mapped genes, and significant results (FDR > 0.01) were visualized using ShinyGO (v0.77).

#### Untargeted Metabolomic Profiling

Global untargeted metabolomic analysis was performed on left ventricular biopsy samples obtained with an 8 mm biopsy punch and flash-frozen. The analysis was performed using ultrahigh performance liquid chromatography-high resolution accurate mass (HRAM) mass spectrometry (HRAM-MS). ^22^ Metabolites were extracted from tissue homogenates spiked with stable isotope-labeled internal standards through protein precipitation using chilled 1:1 methanol: acetonitrile. Ten microliters of the extract were analyzed on a high-resolution accurate-mass (HRAM) platform consisting of Vanquish™ Duo UHPLC system equipped with a dual split sampler configuration, coupled to a Exploris 240 HRAM mass spectrometer (Thermo Fisher Scientific, San Jose, CA, USA). Chromatographic separation was performed in triplicate using hydrophilic interaction liquid chromatography (HILIC) under positive ion mode and Reverse Phase (RP) chromatography under negative ion mode, both at 60°C. HILIC separation was carried out on a Waters XBridge BEH Amide XP column (2.1 mm, 50mm. 2.5 μm) using a gradient elution with mobile phases consisting of 0.2% formic acid in water and acetonitrile. RP separation was performed on a Higgins Targa C18 column (2.1 × 50 mm, 3 μm) with 1mM ammonium acetate in water and acetonitrile as mobile solvents. The HRAM mass spectrometer was operated in Full Scan (120K resolution) acquisition mode and datadependent (DDA) acquisition mode, with 60K resolution in full scan and 7500 ddMS2, both positive and negative polarity modes to acquire the spectral data. Raw data files were acquired through the Xcalibur software (version 4.1, ThermoFisher Scientific, MA, USA) and processed using the Compound Discoverer software (version 3.3.1, ThermoFisher Scientific, Waltham, MA, USA). The workflow used an adaptive curve model with 1 min maximum shift, 10 ppm mass tolerance, and 3 S/N (signal/noise) threshold for retention time alignment. Compound identification was achieved by using mzVault (internal ddMS2 database), mzCloud, and ChemSpider (using formula or exact mass and HMDB, KEGG, LipidMAPS as databases). Similarity searches for all compounds with ddMS2 data was done by using mzCloud and mzLogic algorithms applied to rank order ChemSpider results. For each identified metabolite, the null hypothesis was that there were no differences in area means across the groups tested. Statistical comparisons were performed using Student’s t-test and p-values, along with q-value and fold-change values. Metabolites were classified either as the whole feature list, which includes all the detected signals from the mass spectrometer regardless of whether they have been identified as known metabolites, or as the annotated compounds list, corresponding to known metabolites that have been identified from the whole feature set using computational analysis.

### Supplemental Figures

**Supplemental Figure 1.**
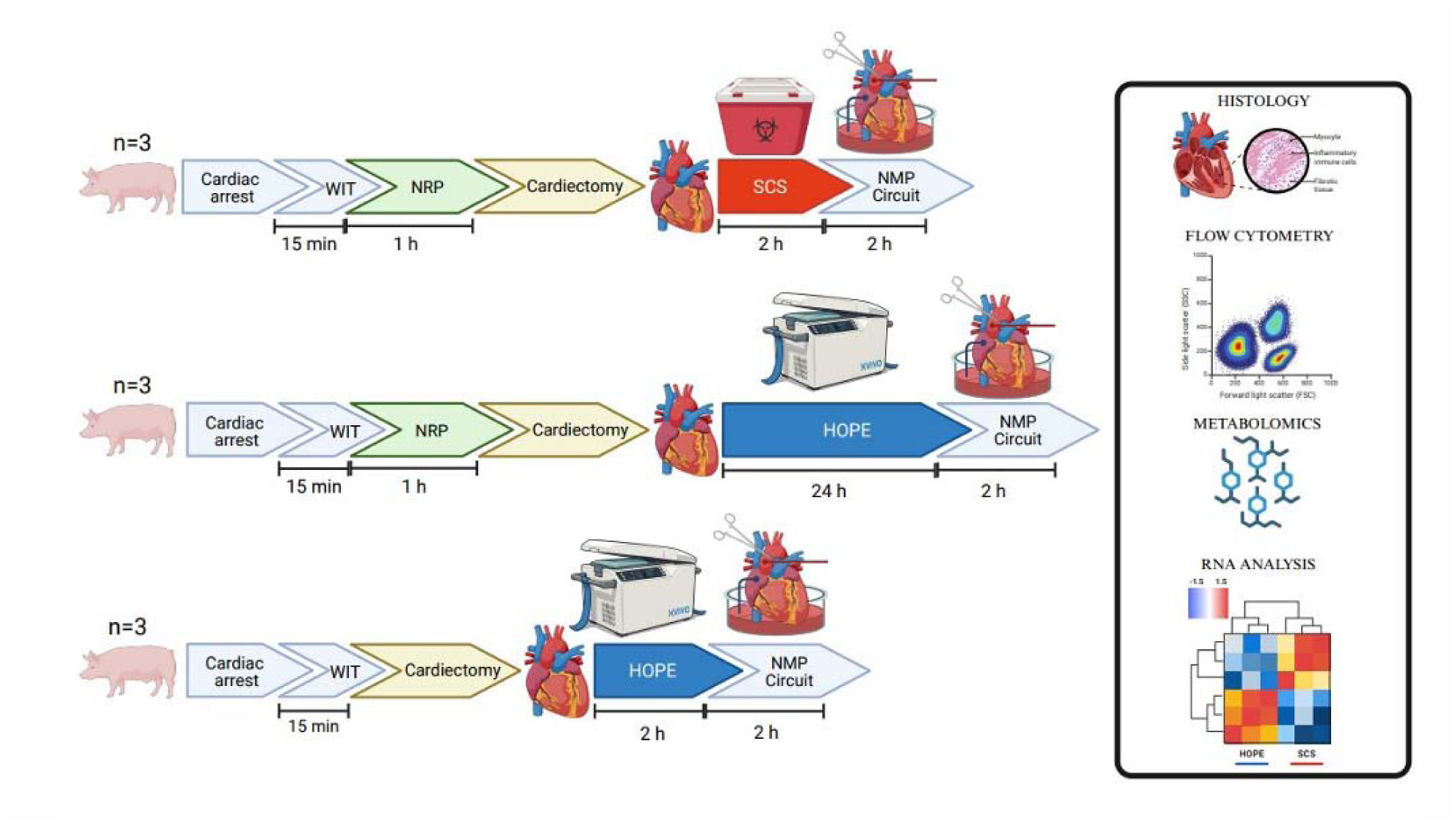
Study design. Schematic representation of the porcine DCD preservation experiments. Nine Yorkshire pigs were divided into three groups (n=3 per group). Top: Following cardiac arrest and 15 min of warm ischemia time (WIT), animals underwent 60 min of normothermic regional perfusion (NRP), cardiectomy, and preservation with static cold storage (SCS) for 2 h before reanimation on a bench-top normothermic machine perfusion (NMP) circuit. Middle: After 15 min WIT and 60 min NRP, hearts were procured and preserved by hypothermic oxygenated perfusion (HOPE) for 24 h prior to NMP reanimation. Bottom: Hearts underwent direct procurement after 15 min WIT without NRP, followed by 2 h of HOPE preservation and subsequent NMP reanimation. Outcomes included histology, flow cytometry, metabolomics, and RNA sequencing to assess cardiomyocyte integrity, molecular signatures, and functional recovery.

**Supplemental Figure 2.**
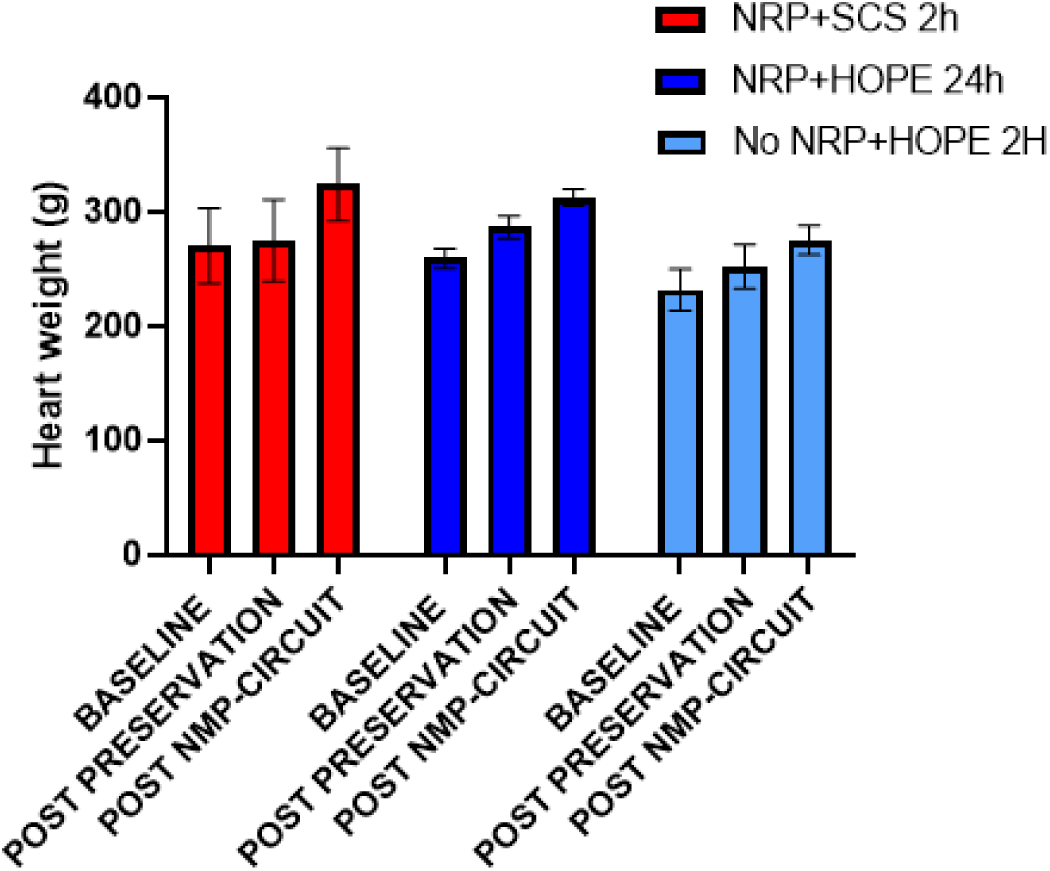
Heart weight changes during preservation and reperfusion. Heart weights were measured at baseline, after preservation, and following 2h of normothermic machine perfusion (NMP). Both NRP + SCS (2h) and NRP + HOPE (24h) hearts showed comparable total weight gain (∼20%), while No NRP + HOPE (2h) hearts exhibited a smaller increase (∼15%). Data are shown as mean ± SEM.

**Supplemental Figure 3.**
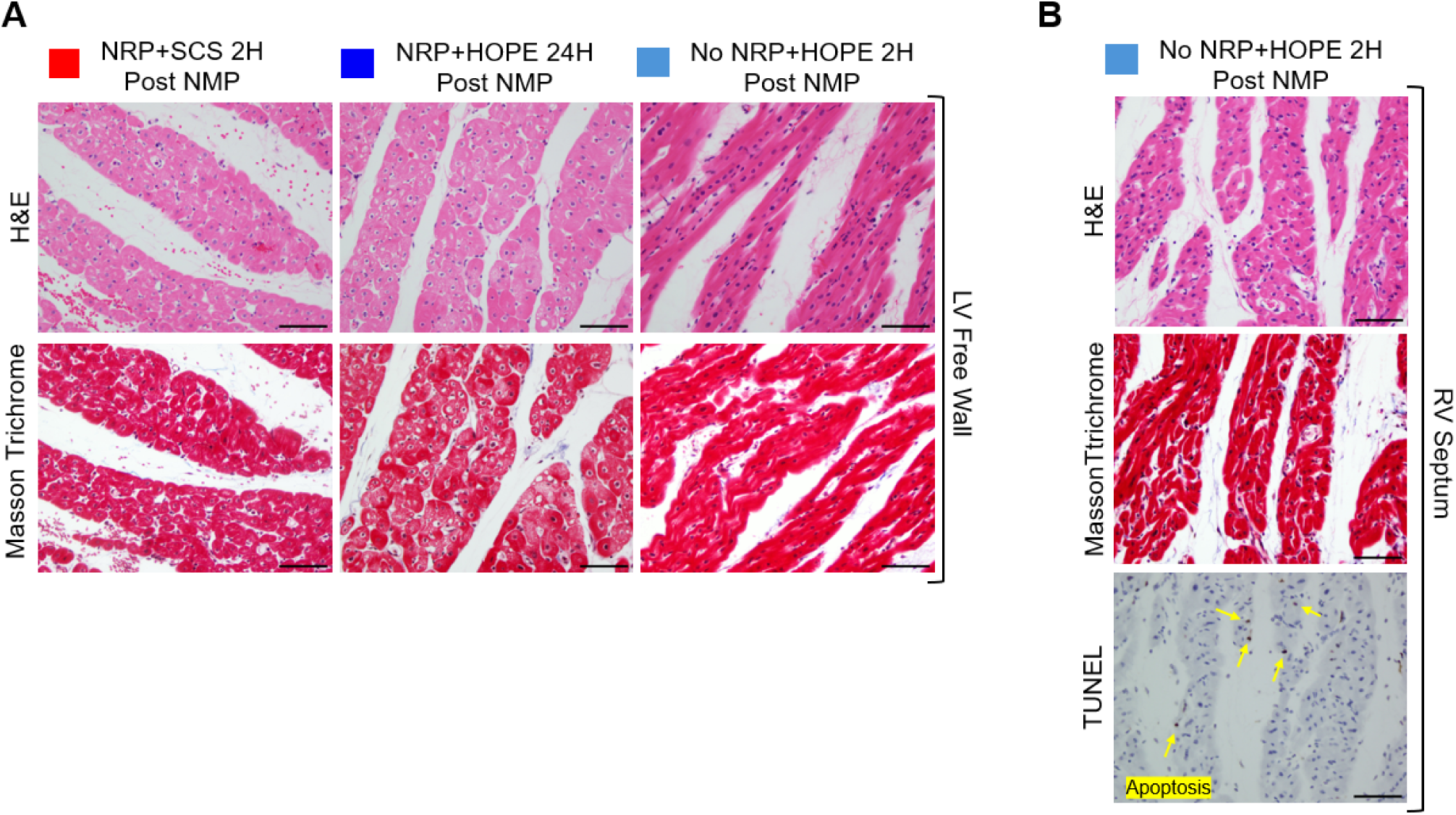
Histological analysis of cardiac tissue. (A) Hematoxylin and Eosin (up) and Masson Trichrome(down) staining of the Left Ventricular Free Wall obtained by endomyocardial biopsies. (B) H&E, Masson Trichrome and TUNEL staining of RV Septum endomyocardial biopsies obtained from a No NRP heart preserved in HOPE for 2 h. Scale bar: 100 µm.

**Supplemental Figure 4.**
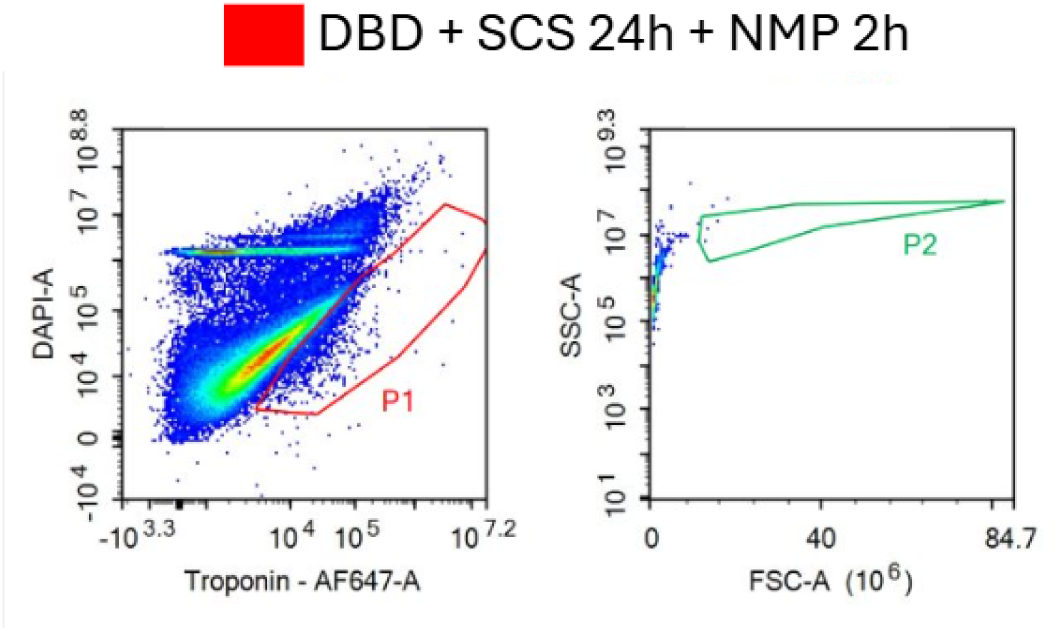
Cardiomyocyte integrity after extended SCS preservation in a model of Donation following Brain Death (DBD). (A) Representative flow cytometry plots of DAPI and troponin staining in isolated cardiomyocytes after DBD procurement followed by 24h SCS and 2h of NMP reanimation circuit. Cardiomyocytes were identified as DAPI-positive, cardiac troponin T-positive events (P1 gate) and further selected for intact cells in a FSC vs SSC plot (P2 gate). The population of isolated intact cardiomyocytes was severely depleted, with no preserved cellular viability following 24h of SCS, despite the more favorable donation model.

**Supplemental Figure 5.**
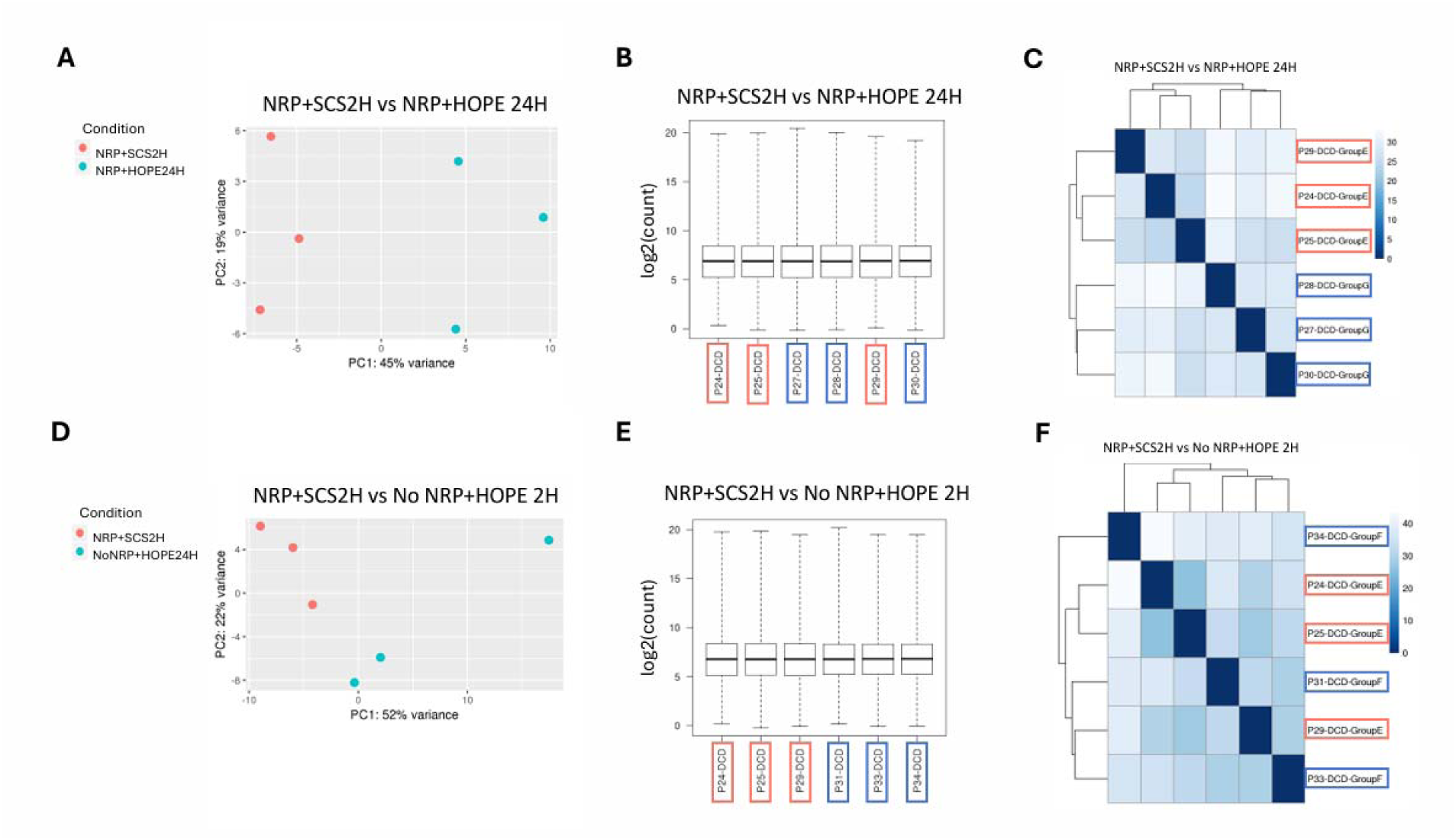
Transcriptomic profiles of DCD hearts preserved with NRP + SCS 2 h versus HOPE 24 h or No NRP + HOPE 2 h. (A–C) Comparison of NRP + SCS 2h and NRP + HOPE 24h (A) Principal component analysis (PCA) shows partial separation between groups. (B) Boxplots of normalized read counts illustrate similar global expression distributions across samples. (C) Unsupervised hierarchical clustering heatmap of the top differentially expressed genes demonstrates partial grouping by preservation strategy, though overall transcriptomic profiles remained similar. (D–F) Comparison of NRP + SCS 2 h and No NRP + HOPE 2 h. (D) PCA again shows modest separation between groups. (E) Boxplots of normalized read counts show comparable expression distributions. (F) Unsupervised clustering heatmap reveals overlap between groups, with no consistent pathway-level enrichment detected.

### Supplemental Videos

- Supplemental Video 1: <u>Circulatory Arrest Swine Open-chest</u>
- Supplemental Video 2: <u>Normothermic Regional Perfusion</u>
- Supplemental Video 3: <u>Bench-Top NMP Reanimation after NRP+SCS 2h</u>
- Supplemental Video 4: <u>Bench-Top NMP Reanimation after NRP+HOPE 24h</u>
- Supplemental Video 5: <u>Bench-Top NMP Reanimation after DBD+SCS 24h</u>
- Supplemental Video 6: <u>Bench-Top NMP Reanimation after No NRP+HOPE 2h</u>

### Supplemental Tables

**Supplemental Table I.**
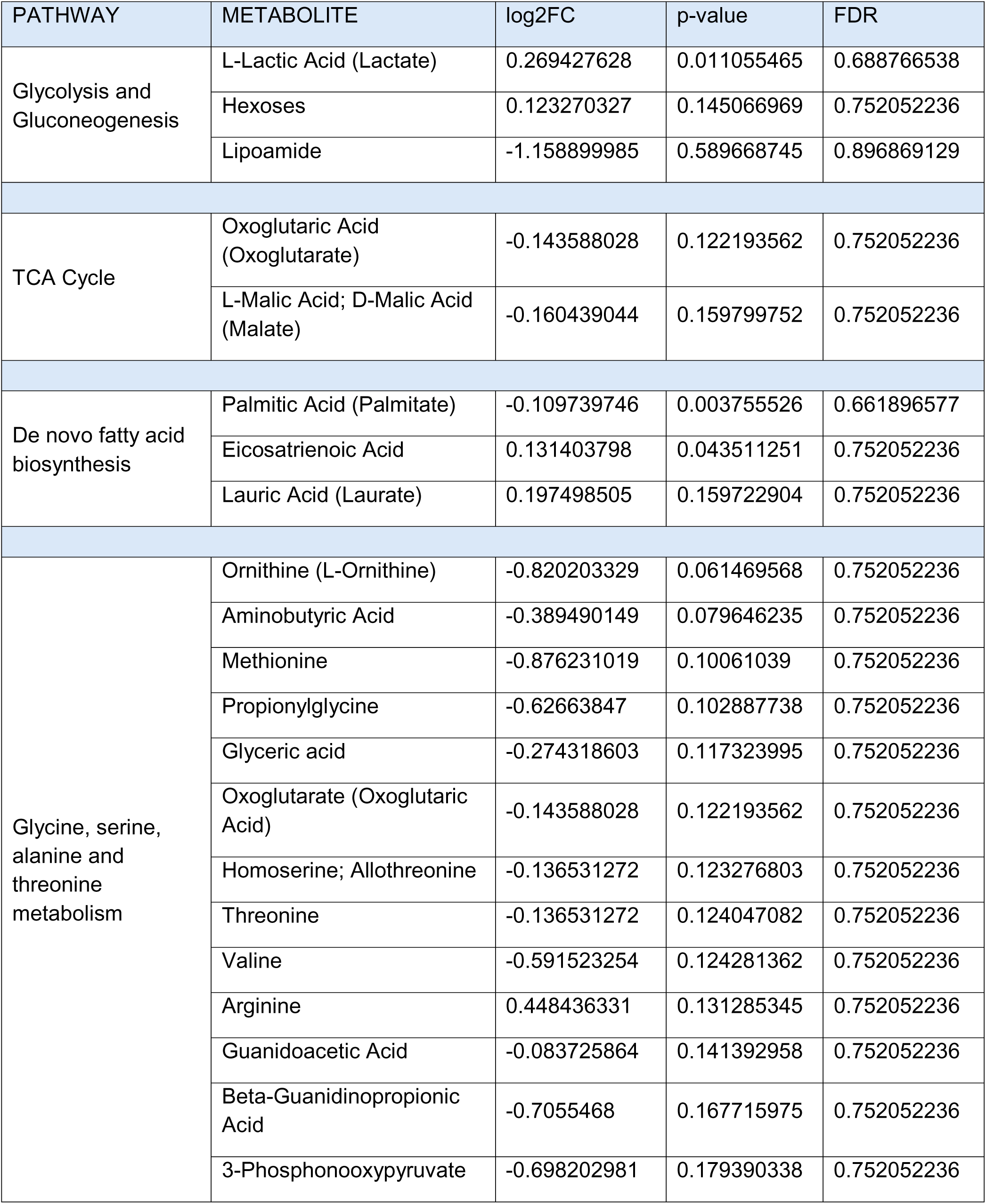
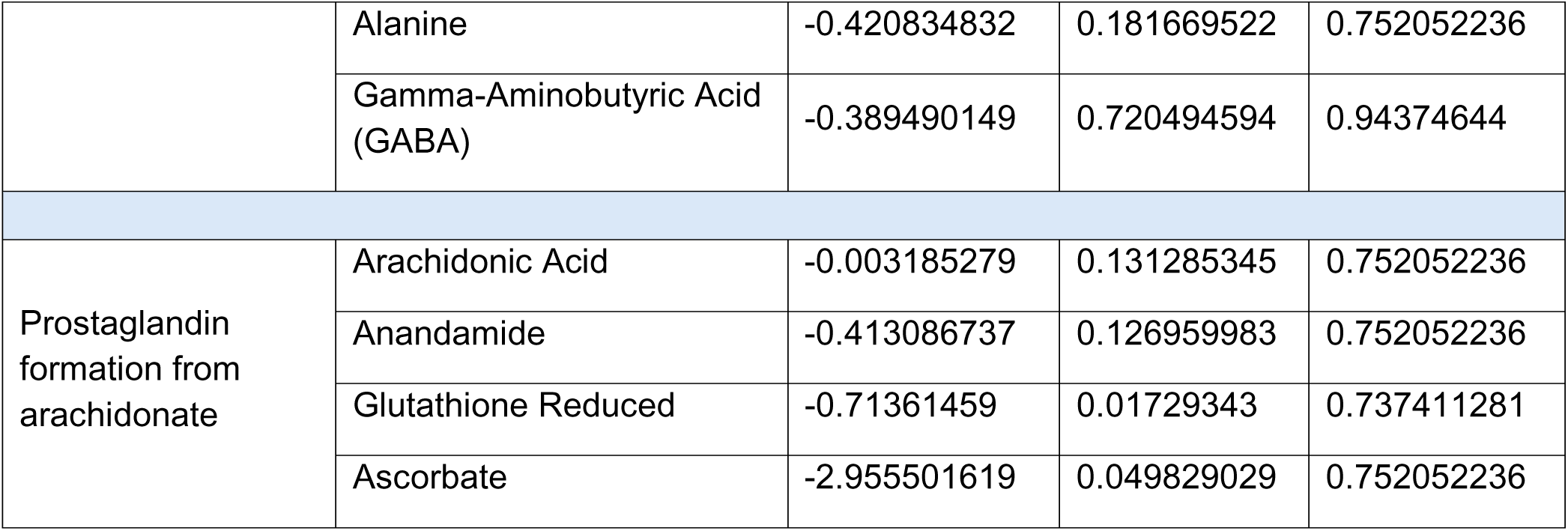
Differentially expressed metabolites between NRP + SCS 2h and NRP + HOPE 24 h hearts associated to the relative functional pathways.

**Supplemental Table II.**
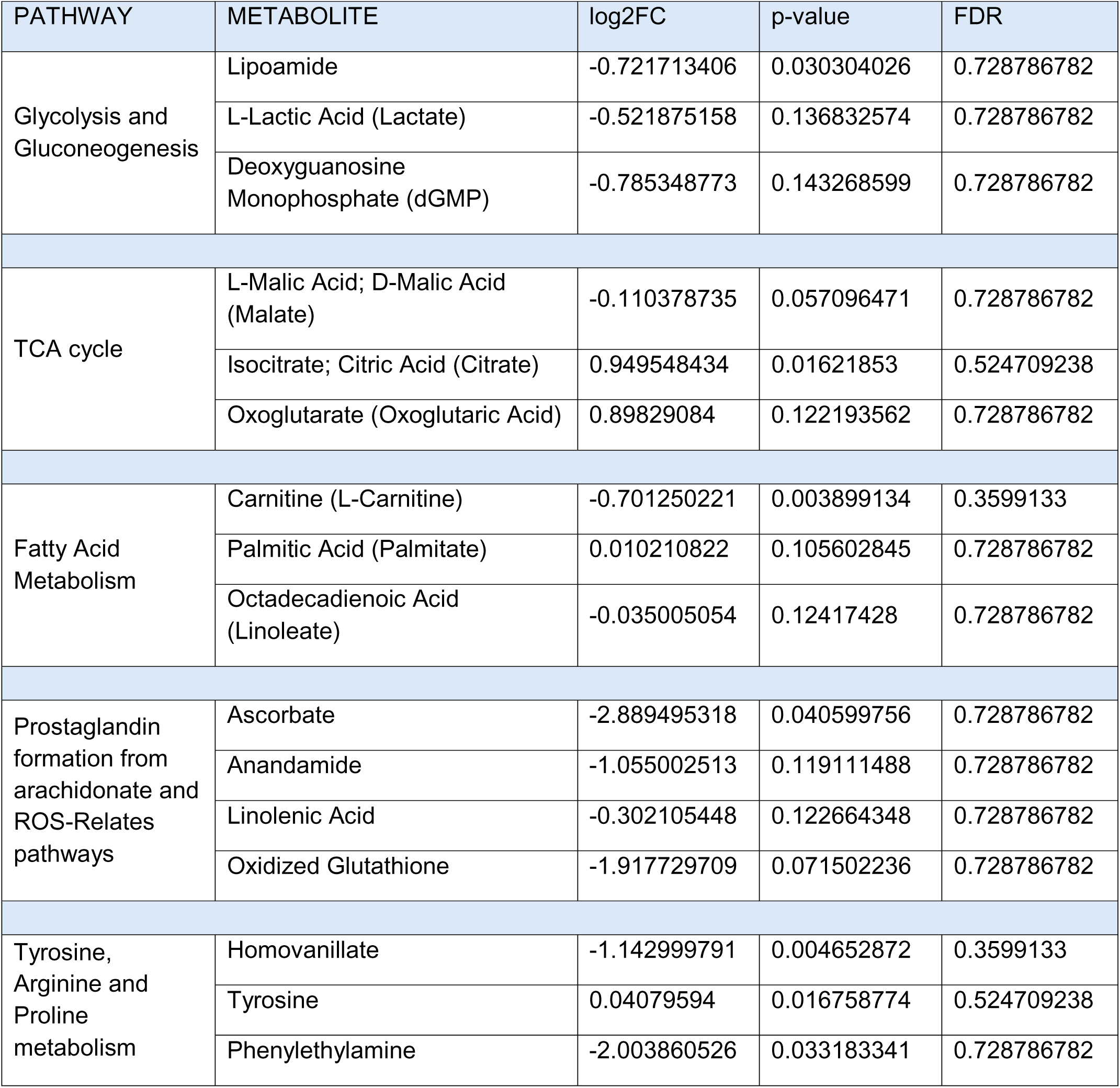
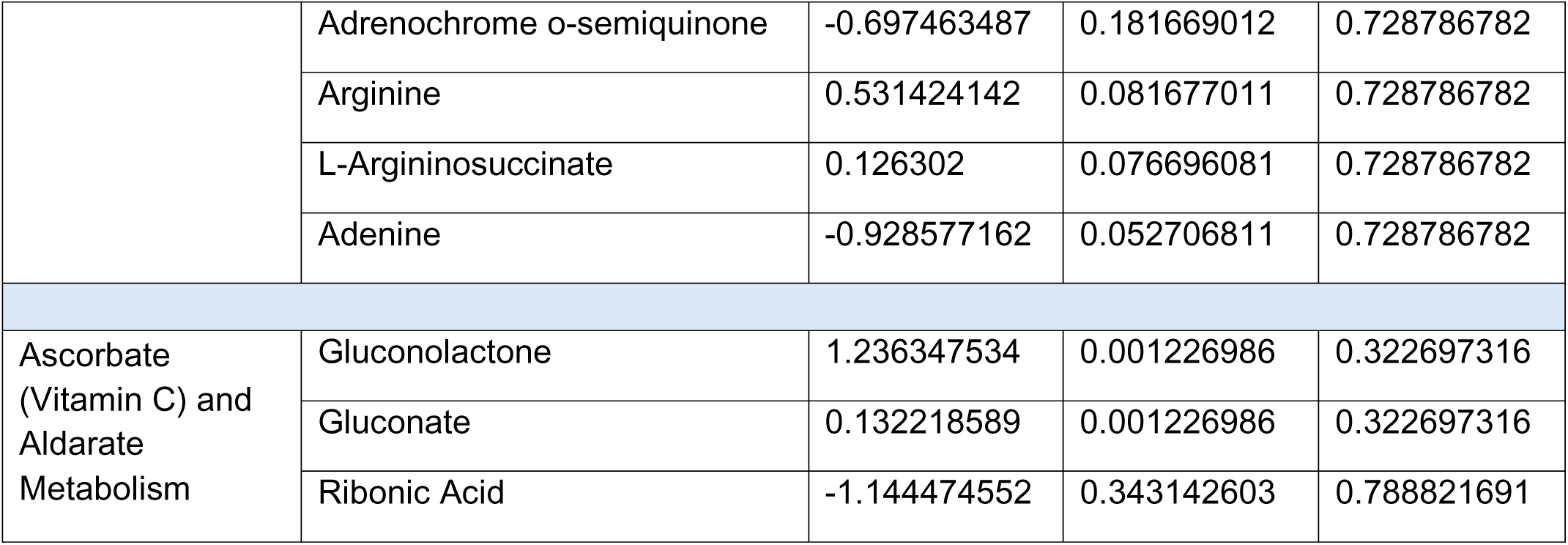
Differentially expressed metabolites between NRP + SCS 2h and No NRP + HOPE 2 h hearts associated to the relative functional pathways.

## References

1. Cameli M, et al., Donor shortage in heart transplantation: How can we overcome this challenge? Front Cardiovasc Med. 2022;9:1001002. doi:10.3389/fcvm.2022.1001002

2. Kittleson MM. Extending the boundaries of organ preservation: hope for heart transplantation. The Lancet. 2024;404(10453):631–633. doi:10.1016/S0140-6736(24)01597-6

3. Coniglio AC, et al., Innovations in Heart Transplantation: A Review. J Card Fail. 2022;28(3):467–476. doi:10.1016/j.cardfail.2021.10.011

4. Goodwin ML, et al., Direct procurement with machine perfusion and normothermic regional perfusion in donation after circulatory death heart transplantation. J Thorac Cardiovasc Surg. 2025;170(1):256–265.e6. doi:10.1016/j.jtcvs.2024.10.033

5. Toro S, Schlendorf KH. Expanding the Donor Pool. Methodist DeBakey Cardiovasc J. 21(3):33–39. doi:10.14797/mdcvj.1577

6. Tessari C, et al., Ex-Vivo Heart Perfusion Machines in DCD Heart Transplantation Model: The State of Art. Transpl Int. 2025;38:12987. doi:10.3389/ti.2025.12987

7. Finotti M, et al., The Dallas Donation after Circulatory Death Transplantation Summit: expanding donation after circulatory death procedures through process improvement, broader utilization, and innovation. Hepatobiliary Surg Nutr. 2024;13(5):824–836. doi:10.21037/hbsn-23-503

8. Arnold M, et al., Metabolic Considerations in Direct Procurement and Perfusion Protocols with DCD Heart Transplantation. Int J Mol Sci. 2024;25(8):4153. doi:10.3390/ijms25084153

9. Joshi Y, et al., The Rapidly Evolving Landscape of DCD Heart Transplantation. Curr Cardiol Rep. 2024;26(12):1499–1507. doi:10.1007/s11886-024-02148-w

10. Gouchoe DA, et al.,Rise of the machines: Normothermic regional perfusion use in heart transplantation in the United States. J Thorac Cardiovasc Surg. 2025;170(1):268–276.e12. doi:10.1016/j.jtcvs.2024.12.001

11. Kucera JA, et al., On-Table Reanimation of a Pediatric Heart from Donation after Circulatory Death. N Engl J Med. 2025;393(3):275–280. doi:10.1056/NEJMoa2503487

12. Vervoorn MT, et al., A novel cardioprotective perfusion protocol prevents functional decline during extended normothermic ex situ heart perfusion of marginal porcine hearts. J Heart Lung Transplant. 2025;44(6):961–971. doi:10.1016/j.healun.2024.10.016

13. Schuster A, et al., An isolated perfused pig heart model for the development, validation and translation of novel cardiovascular magnetic resonance techniques. J Cardiovasc Magn Reson. 2010;12(1):53. doi:10.1186/1532-429X-12-53

14. Rosenstrauch D, et al., Ex Vivo Resuscitation of Adult Pig Hearts. Tex Heart Inst J. 2003;30(2):121–127.

15. Colah S, et al.,Ex vivo perfusion of the swine heart as a method for pre-transplant assessment. Perfusion. 2012;27(5):408–413. doi:10.1177/0267659112449035

16. Ughetto A, et al.,Heart graft preservation technics and limits: an update and perspectives. Front Cardiovasc Med. 2023;10:1248606. doi:10.3389/fcvm.2023.1248606

17. Brouckaert J, et al.,*v*Non-ischaemic preservation of the donor heart in heart transplantation: protocol design and rationale for a randomised, controlled, multicentre clinical trial across eight European countries. BMJ Open. 2023;13(12):e073729. doi:10.1136/bmjopen-2023-073729

18. Paez JR, et al., Investigating Cardiac Temperature During Heart Transplantation Using the Static Cold Storage Paradigm. Transplantation. 2025;109(3):e148. doi:10.1097/TP.0000000000005185

19. McGiffin et al.,Hypothermic oxygenated perfusion (HOPE) safely and effectively extends acceptable donor heart preservation times: Results of the Australian and New Zealand trial. J Heart Lung Transplant. 2024;43(3):485–495. doi:10.1016/j.healun.2023.10.020

20. Camillo C, et al., Ex Vivo Hypothermic Perfusion Enables 48-Hour Heart Preservation and Bench-Top Functional Recovery via Normothermic Reperfusion in a Porcine Model. medRxiv. Preprint posted online September 5, 2025:2025.09.03.25335053. doi:10.1101/2025.09.03.25335053

