## Supplemental Material for "Ex vivo hypothermic oxygenated perfusion allows extended heart preservation in a donation-after-circulatory-death porcine model"

**Supplemental Figures**

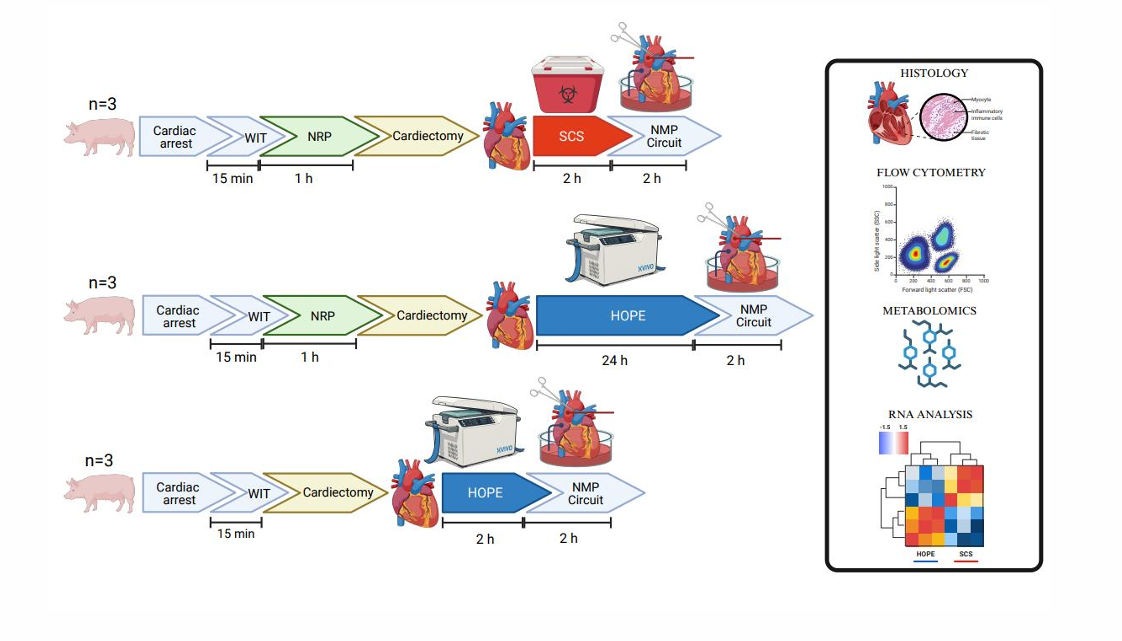
**Supplemental Figure 1. Study design.** Schematic representation of the porcine DCD preservation experiments. Nine Yorkshire pigs were divided into three groups (n=3 per group). Top: Following cardiac arrest and 15 min of warm ischemia time (WIT), animals underwent 60 min of normothermic regional perfusion (NRP), cardiectomy, and preservation with static cold storage (SCS) for 2 h before reanimation on a bench-top normothermic machine perfusion (NMP) circuit. Middle: After 15 min WIT and 60 min NRP, hearts were procured and preserved by hypothermic oxygenated perfusion (HOPE) for 24 h prior to NMP reanimation. Bottom: Hearts underwent direct procurement after 15 min WIT without NRP, followed by 2 h of HOPE preservation and subsequent NMP reanimation. Outcomes included histology, flow cytometry, metabolomics, and RNA sequencing to assess cardiomyocyte integrity, molecular signatures, and functional recovery.

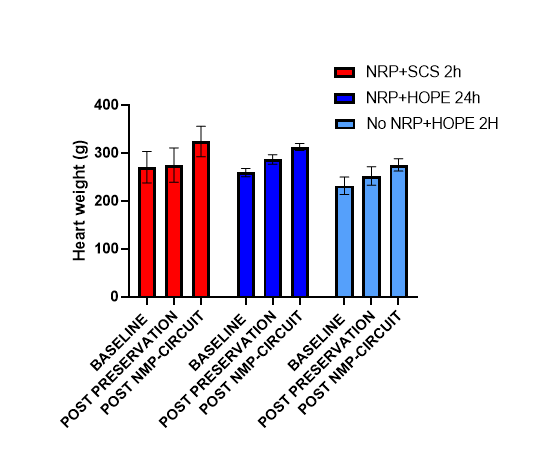

**Supplemental Figure 2. Heart weight changes during preservation and reperfusion.**
Heart weights were measured at baseline, after preservation, and following 2h of normothermic machine perfusion (NMP). Both NRP + SCS (2h) and NRP + HOPE (24h) hearts showed comparable total weight gain (~20%), while No NRP + HOPE (2h) hearts exhibited a smaller increase (~15%). Data are shown as mean ± SEM.

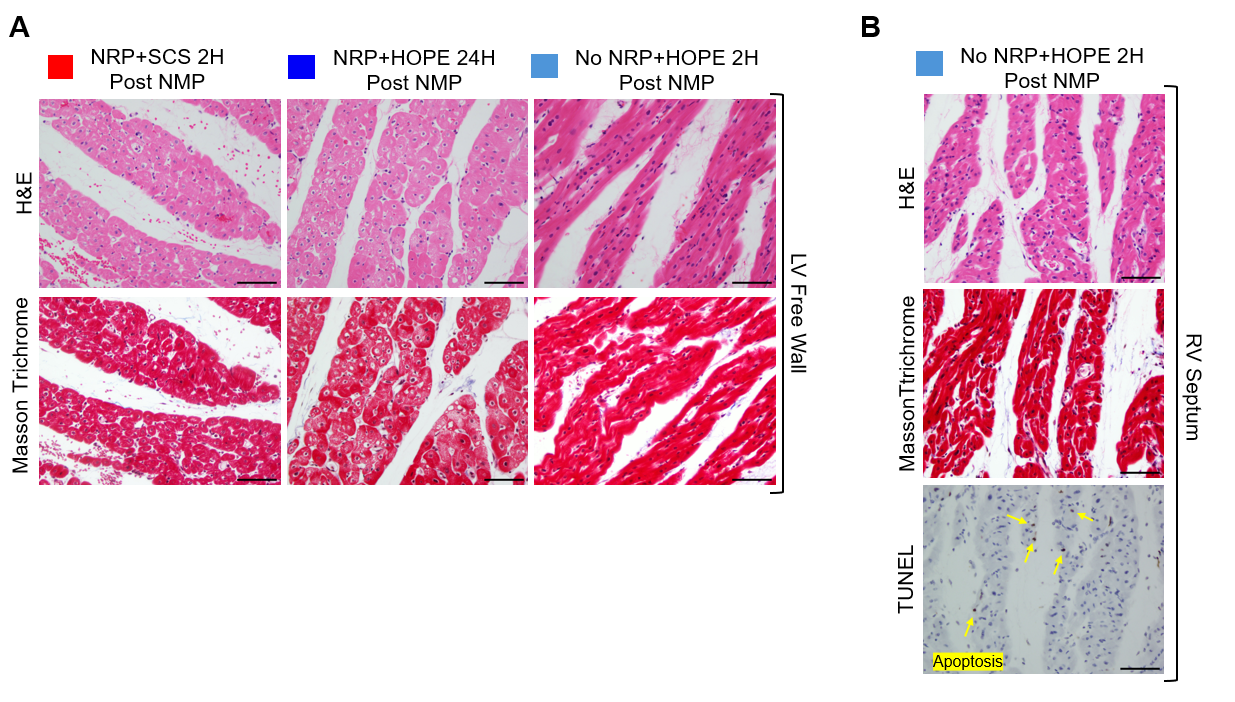

**Supplemental Figure 3. Histological analysis of cardiac tissue.** (A) Hematoxylin and Eosin (up) and Masson Trichrome(down) staining of the Left Ventricular Free Wall obtained by endomyocardial biopsies. (B) H&E, Masson Trichrome and TUNEL staining of RV Septum endomyocardial biopsies obtained from a No NRP heart preserved in HOPE for 2 h. Scale bar: 100 µm.

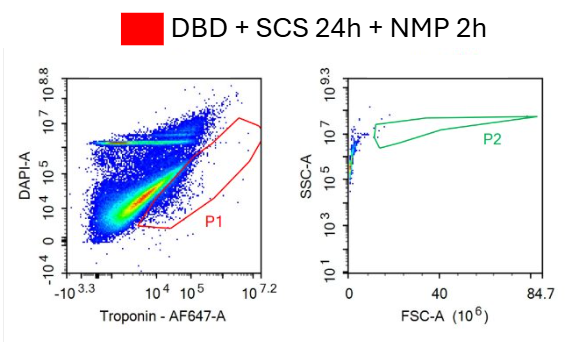

**Supplemental Figure 4. Cardiomyocyte integrity after extended SCS preservation in a model of Donation following Brain Death (DBD).** (A) Representative flow cytometry plots of DAPI and troponin staining in isolated cardiomyocytes after DBD procurement followed by 24h SCS and 2h of NMP reanimation circuit. Cardiomyocytes were identified as DAPI-positive, cardiac troponin T- positive events (P1 gate) and further selected for intact cells in a FSC vs SSC plot (P2 gate). The population of isolated intact cardiomyocytes was severely depleted, with no preserved cellular viability following 24h of SCS, despite the more favorable donation model.

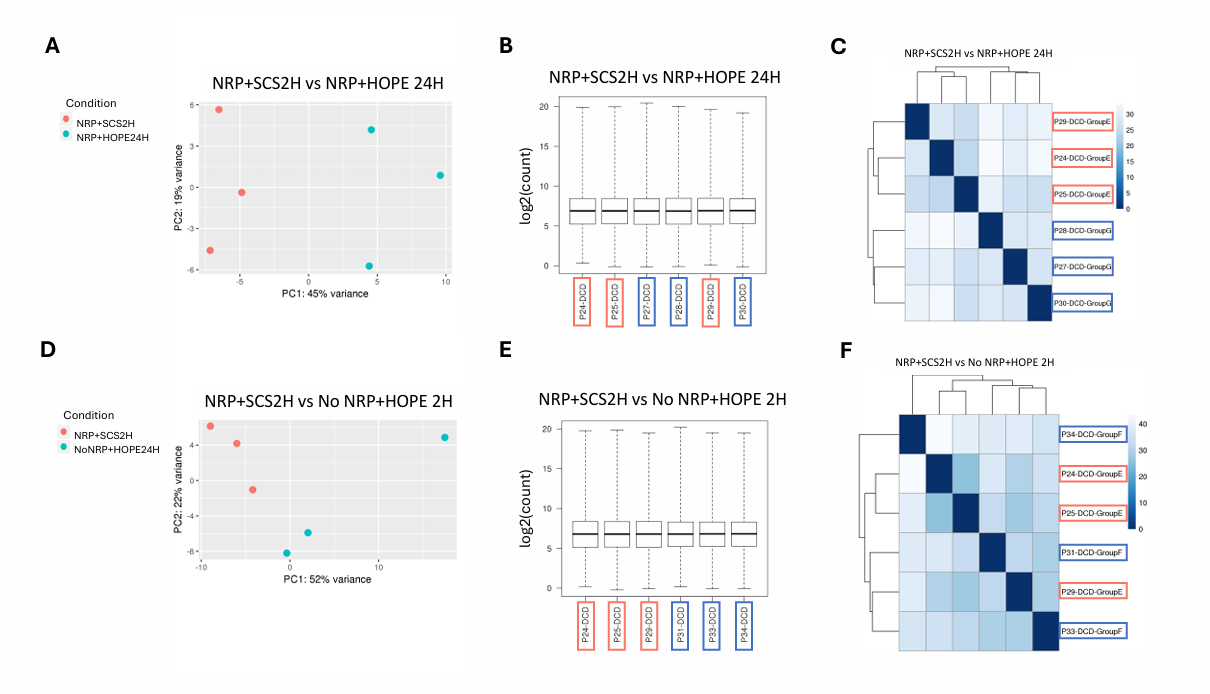
**Supplemental Figure 5. Transcriptomic profiles of DCD hearts preserved with NRP + SCS 2 h versus HOPE 24 h or No NRP + HOPE 2 h.** (A–C) Comparison of NRP + SCS 2h and NRP + HOPE 24h (A) Principal component analysis (PCA) shows partial separation between groups. (B) Boxplots of normalized read counts illustrate similar global expression distributions across samples. (C) Unsupervised hierarchical clustering heatmap of the top differentially expressed genes demonstrates partial grouping by preservation strategy, though overall transcriptomic profiles remained similar. (D–F) Comparison of NRP + SCS 2 h and No NRP + HOPE 2 h. (D) PCA again shows modest separation between groups. (E) Boxplots of normalized read counts show comparable expression distributions. (F) Unsupervised clustering heatmap reveals overlap between groups, with no consistent pathway-level enrichment detected.

**Supplemental Videos**

- **Supplemental Video 1:** [Circulatory Arrest Swine Open-chest](https://drive.google.com/file/d/1P6LTgNNpRRfvwuu5v8fna5DP4pqVOAWB/view?usp=drive_link)
- **Supplemental Video 2:** [Normothermic Regional Perfusion](https://drive.google.com/file/d/1w7-hBop79pBBinpfgEkW0mGywKB2I7yr/view?usp=drive_link)
- **Supplemental Video 3:** [Bench-Top NMP Reanimation after NRP+SCS 2h](https://drive.google.com/file/d/1PQXTt5tRN_-z-Lw-uqPYOCCno0_DjoSN/view?usp=drive_link)
- **Supplemental Video 4:** [Bench-Top NMP Reanimation after NRP+HOPE 24h](https://drive.google.com/file/d/1rRygT-WdkTnWvxhwNYjZojVp4hmXbSpT/view?usp=drive_link)
- **Supplemental Video 5:** [Bench-Top NMP Reanimation after DBD+SCS 24h](https://drive.google.com/file/d/1A8a2LEG_vfrkXbbJwbEsEBogWF61XmvF/view?usp=drive_link)
- **Supplemental Video 6:** [Bench-Top NMP Reanimation after No NRP+HOPE 2h](https://drive.google.com/file/d/1Gomi78I8ezdHxea4HVBZMwbY7yKDZIav/view?usp=drive_link)

**Supplemental Tables.**

| **PATHWAY** | **METABOLITE** | **log2FC** | **p-value** | **FDR** |
| --- | --- | --- | --- | --- |
| Glycolysis and Gluconeogenesis | L-Lactic Acid (Lactate) | 0.269427628 | 0.011055465 | 0.688766538 |
|  | Hexoses | 0.123270327 | 0.145066969 | 0.752052236 |
|  | Lipoamide | -1.158899985 | 0.589668745 | 0.896869129 |
| TCA Cycle | Oxoglutaric Acid (Oxoglutarate) | -0.143588028 | 0.122193562 | 0.752052236 |
|  | L-Malic Acid; D-Malic Acid (Malate) | -0.160439044 | 0.159799752 | 0.752052236 |
| De novo fatty acid biosynthesis | Palmitic Acid (Palmitate) | -0.109739746 | 0.003755526 | 0.661896577 |
|  | Eicosatrienoic Acid | 0.131403798 | 0.043511251 | 0.752052236 |
|  | Lauric Acid (Laurate) | 0.197498505 | 0.159722904 | 0.752052236 |
| Glycine, serine, alanine and threonine metabolism | Ornithine (L-Ornithine) | -0.820203329 | 0.061469568 | 0.752052236 |
|  | Aminobutyric Acid | -0.389490149 | 0.079646235 | 0.752052236 |
|  | Methionine | -0.876231019 | 0.10061039 | 0.752052236 |
|  | Propionylglycine | -0.62663847 | 0.102887738 | 0.752052236 |
|  | Glyceric acid | -0.274318603 | 0.117323995 | 0.752052236 |
|  | Oxoglutarate (Oxoglutaric Acid) | -0.143588028 | 0.122193562 | 0.752052236 |
|  | Homoserine; Allothreonine | -0.136531272 | 0.123276803 | 0.752052236 |
|  | Threonine | -0.136531272 | 0.124047082 | 0.752052236 |
|  | Valine | -0.591523254 | 0.124281362 | 0.752052236 |
|  | Arginine | 0.448436331 | 0.131285345 | 0.752052236 |
|  | Guanidoacetic Acid | -0.083725864 | 0.141392958 | 0.752052236 |
|  | Beta-Guanidinopropionic Acid | -0.7055468 | 0.167715975 | 0.752052236 |
|  | 3-Phosphonooxypyruvate | -0.698202981 | 0.179390338 | 0.752052236 |
|  | Alanine | -0.420834832 | 0.181669522 | 0.752052236 |
|  | Gamma-Aminobutyric Acid (GABA) | -0.389490149 | 0.720494594 | 0.94374644 |
| Prostaglandin formation from arachidonate | Arachidonic Acid | -0.003185279 | 0.131285345 | 0.752052236 |
|  | Anandamide | -0.413086737 | 0.126959983 | 0.752052236 |
|  | Glutathione Reduced | -0.71361459 | 0.01729343 | 0.737411281 |
|  | Ascorbate | -2.955501619 | 0.049829029 | 0.752052236 |

**Supplemental Table I.** Differentially expressed metabolites between NRP + SCS 2h and NRP + HOPE 24 h hearts associated to the relative functional pathways.

| **PATHWAY** | **METABOLITE** | **log2FC** | **p-value** | **FDR** |
| --- | --- | --- | --- | --- |
| Glycolysis and Gluconeogenesis | Lipoamide | -0.721713406 | 0.030304026 | 0.728786782 |
|  | L-Lactic Acid (Lactate) | -0.521875158 | 0.136832574 | 0.728786782 |
|  | Deoxyguanosine Monophosphate (dGMP) | -0.785348773 | 0.143268599 | 0.728786782 |
| TCA cycle | L-Malic Acid; D-Malic Acid (Malate) | -0.110378735 | 0.057096471 | 0.728786782 |
|  | Isocitrate; Citric Acid (Citrate) | 0.949548434 | 0.01621853 | 0.524709238 |
|  | Oxoglutarate (Oxoglutaric Acid) | 0.89829084 | 0.122193562 | 0.728786782 |
| Fatty Acid Metabolism | Carnitine (L-Carnitine) | -0.701250221 | 0.003899134 | 0.3599133 |
|  | Palmitic Acid (Palmitate) | 0.010210822 | 0.105602845 | 0.728786782 |
|  | Octadecadienoic Acid (Linoleate) | -0.035005054 | 0.12417428 | 0.728786782 |
| Prostaglandin formation from arachidonate and ROS-Relates pathways | Ascorbate | -2.889495318 | 0.040599756 | 0.728786782 |
|  | Anandamide | -1.055002513 | 0.119111488 | 0.728786782 |
|  | Linolenic Acid | -0.302105448 | 0.122664348 | 0.728786782 |
|  | Oxidized Glutathione | -1.917729709 | 0.071502236 | 0.728786782 |
| Tyrosine, Arginine and Proline metabolism | Homovanillate | -1.142999791 | 0.004652872 | 0.3599133 |
|  | Tyrosine | 0.04079594 | 0.016758774 | 0.524709238 |
|  | Phenylethylamine | -2.003860526 | 0.033183341 | 0.728786782 |
|  | Adrenochrome o-semiquinone | -0.697463487 | 0.181669012 | 0.728786782 |
|  | Arginine | 0.531424142 | 0.081677011 | 0.728786782 |
|  | L-Argininosuccinate | 0.126302 | 0.076696081 | 0.728786782 |
|  | Adenine | -0.928577162 | 0.052706811 | 0.728786782 |
| Ascorbate (Vitamin C) and Aldarate Metabolism | Gluconolactone | 1.236347534 | 0.001226986 | 0.322697316 |
|  | Gluconate | 0.132218589 | 0.001226986 | 0.322697316 |
|  | Ribonic Acid | -1.144474552 | 0.343142603 | 0.788821691 |

**Supplemental Table II**. Differentially expressed metabolites between NRP + SCS 2h and No NRP + HOPE 2 h hearts associated to the relative functional pathways.
